# Assessing the performance of GPT-4 in the filed of osteoarthritis and orthopaedic case consultation

**DOI:** 10.1101/2023.08.06.23293735

**Authors:** Juntan Li, Xiang Gao, Tianxu Dou, Yuyang Gao, Wannan Zhu

## Abstract

**Background:** Large Language Models (LLMs) like GPT-4 demonstrate potential applications in diverse areas, including healthcare and patient education. This study evaluates GPT-4’s competency against osteoarthritis (OA) treatment guidelines from the United States and China and assesses its ability in diagnosing and treating orthopedic diseases.

**Methods:** Data sources included OA management guidelines and orthopedic examination case questions. Queries were directed to GPT-4 based on these resources, and its responses were compared with the established guidelines and cases. The accuracy and completeness of GPT-4’s responses were evaluated using Likert scales, while case inquiries were stratified into four tiers of correctness and completeness.

**Results:** GPT-4 exhibited strong performance in providing accurate and complete responses to OA management recommendations from both the American and Chinese guidelines, with high Likert scale scores for accuracy and completeness. It demonstrated proficiency in handling clinical cases, making accurate diagnoses, suggesting appropriate tests, and proposing treatment plans. Few errors were noted in specific complex cases.

**Conclusions:** GPT-4 exhibits potential as an auxiliary tool in orthopedic clinical practice and patient education, demonstrating high accuracy and completeness in interpreting OA treatment guidelines and analyzing clinical cases. Further validation of its capabilities in real-world clinical scenarios is needed.

## Instruction

Large Language Models (LLMs) delineate a category of machine learning algorithms designed to generate text resembling human-like semantic and syntactic structures. These models, trained on expansive collections of internet-derived text, exploit algorithms originating from the transformer architecture, such as the Generative Pretrained Transformer (GPT) series, pioneered by OpenAI[1]. Leveraging patterns discerned during the training phase, these models interpret contextual input and estimate the subsequent word in an ongoing sentence[2–3]. LLMs have demonstrated substantial promise across a wide spectrum of applications. A prominent example is ChatGPT, which manifests impressive human-like expressiveness and reasoning capabilities. Its use cases span an array of tasks, including drafting emails, crafting code, creative writing and even translating intricate medical lexicon into simple language comprehensible for laymen[4–5]. Further, it has been employed as a preparatory tool for medical board examinations, thus evidencing its immense potential in education[6–7].

GPT-4, as the most recent version in the Generative Pretrained Transformer series initiated by OpenAI, constitutes a notable progress in the sphere of LLMs[8–9]. As opposed to its forerunner, GPT-4 has displayed an enhanced competency in numerous tasks[10–11]. Research outcomes have indicated that GPT-4 surpasses ChatGPT in simulations pertaining to medical board examinations, as it demonstrates higher precision and superior comprehension of intricate, high-level queries. This infers enhanced abilities on part of GPT-4 in context comprehension and problem resolution[12]. In addition, GPT-4 has exhibited significant enhancements in converting sophisticated medical terminologies into layman’s language, thereby presenting potential applications in educating patients and facilitating healthcare communication[13]. Despite these remarkable skills, it is crucial to acknowledge that LLMs, inclusive of GPT-4, do not interpret text as humans do. They lack consciousness and any statement they generate regarding the world necessitates fact-checking for accuracy. Hence, it is conceivable that the model may yield incorrect information owing to its inherent “illusions”.

Osteoarthritis (OA) is a chronic degenerative joint disease that poses a significant public health challenge due to its high prevalence and disability rate[14–15]. Globally, it impacts hundreds of millions of individuals, with the incidence increasing with age and being more prevalent in women than men[16]. Consequently, OA is one of the leading causes of global disability[17]. The clinical manifestations of OA include joint pain, stiffness, and loss of function, primarily affecting the knees, hands, hip, and spine[18]. These symptoms range from mild to severe, often limiting daily activities and decreasing the quality of life of affected individuals. Multiple treatment options exist for OA, ranging from non-pharmacological interventions such as physical therapy and lifestyle modifications, to pharmacological treatments and surgical procedures[19]. Each of these treatment modalities carries associated costs, contributing to a substantial economic burden related to OA management[20]. This burden encompasses both direct medical expenses (e.g., consultations, medications, hospitalizations, surgeries) and indirect costs such as productivity losses due to disability or premature death. In addition, the importance of self-education in OA management cannot be overstated[21]. Patients who are well-informed about their condition are more likely to actively participate in their care, adhere to prescribed treatment regimens, and achieve improved health outcomes.

In light of this, the present study aims to explore the potential of GPT4 in the field of OA. We aim to evaluate the accuracy and completeness of GPT-4 responses compared to established treatment guidelines in China and the United States. A key objective is to assess the feasibility of using GPT-4 as a tool for patient health education, serving as an assistant to clinicians. Moreover, we strive to understand the performance of GPT-4 in diagnosing and treating orthopedic diseases.

## Materials and Methods

### Data source

The present study incorporates a broad array of data sources encompassing: (1) the Evidence-Based Clinical Practice Guideline for the Management of Osteoarthritis of the Knee (Non-Arthroplasty), promulgated by the American Academy of Orthopaedic Surgery (AAOS) in 2021, which constitutes 28 OA management recommendations arranged hierarchically into four-star categories utilizing visualization techniques[22]. (2) The 2021 Guidelines for Osteoarthritis Diagnosis and Treatment issued by the Chinese Orthopedic Association, which furnishes 30 propositions addressing a set of 15 paramount clinical concerns identified among orthopedic physicians. These propositions are discretely assigned into A, B, and C tiers based on their respective recommendation levels[30]. (3) An assortment of fifty case analysis examination questions randomly selected from the question repository of the Chinese Orthopaedic Specialist Examination.

### GPT4 prompt and response generation

A prompt serves as the crucial steering wheel in language models, dictating the direction of the generated response and significantly impacting the quality, relevance, and safety of the model’s output. The quality and nature of the output produced by GPT-4 are significantly influenced by the prompt provided.

Within the context of the AAOS guidelines, GPT-4 is directly interrogated based on the specifics of these recommendations. An exemplar query could be, “ Are canes recommended for improving function and quality of life for osteoarthritis patients ?” Considering the formidable reasoning and logical capabilities of GPT-4, we further probe, “Given that a 4-star rating represents the pinnacle of recommendation, how many stars would you accord this particular recommendation?” Subsequently, the responses generated by GPT-4 are compared with the established guidelines for comparison. In relation to the Chinese osteoarthritis guidelines, GPT-4 was directly queried using the 15 clinically pertinent questions outlined within these guidelines. The subsequent analysis focused on delineating the differences between GPT-4’s responses and the recommendations explicitly enumerated in the guidelines. With respect to testing the case inquiry abilities, we initially provide case information, after which GPT-4 is assigned to respond to these cases concerning further radiological examinations, primary diagnoses, and therapeutic strategies. This procedure is intended to assess its potential effectiveness as an adept assistant in the field of orthopedic.

### Evaluation of response

In terms of recommendation index prediction, directly compare the recommended scores predicted by GPT-4 with those of AAOS. When comparing with the guideline recommendations, two independent evaluators (Juntan Li and Xiang Gao) assessed the accuracy and completeness of GPT-4 responses. If there is a discrepancy in the evaluation, a third researcher is brought in for discussion to determine the final ranking. A 5-point Likert scale was employed to appraise accuracy (5 = correct, 4 = more correct than incorrect, 3= approximately equal correct and incorrect, 2 = more incorrect than correct, 1 = completely incorrect). The degree of completeness was evaluated using a 3-point Likert scale (3 = comprehensive, 2 = adequate, 1 = incomplete). Regarding case inquiries, the responses generated by GPT-4 are classified into four tiers: 4 = comprehensive, 3 =correct but inadequate, 2 =mixed with correct and incorrect/outdated data, and 1 = completely incorrect. This stratified classification serves to evaluate the proficiency of GPT-4 in discerning orthopedic pathologies.

### Statistical analysis

The comparative results are archived in Excel, facilitating the computation of the proportionate distribution of GPT-4 responses at each level. A descriptive statistical analysis is subsequently executed, and data visualization is achieved using the coding interpreter of GPT-4.

## Results

### 1.1 AAOS guideline

In the AAOS guidelines, recommendations related to OA are ranked from 1 to 4, and GPT-4 also assigns ratings to recommendations on a similar scale of 1-4(Table 1 and Supplementary Table 1). Occasionally, GPT-4 may provide a neutral rating, such as 2 or 3. In such instances, we categorize it as ’largely correct’. If the ratings completely match, they are deemed ’correct’, while completely different ratings are labeled ’incorrect’. As depicted in Figure 1A, the correct match is at 46.4%, while largely correct ratings account for 50%. Only a small fraction of 3.6% are incorrect, demonstrating GPT-4’s superior performance in rating OA recommendations. Figure 1B presents a confusion matrix comparing guideline rankings with those predicted by GPT-4. When neutral responses occur, we include instances where the ratings coincide in the figure. It can be observed that GPT-4 exhibits impressive accuracy in the delineation of recommendation levels, regardless of whether they belong to the 1-4 ranking range.

**Figure 1:**
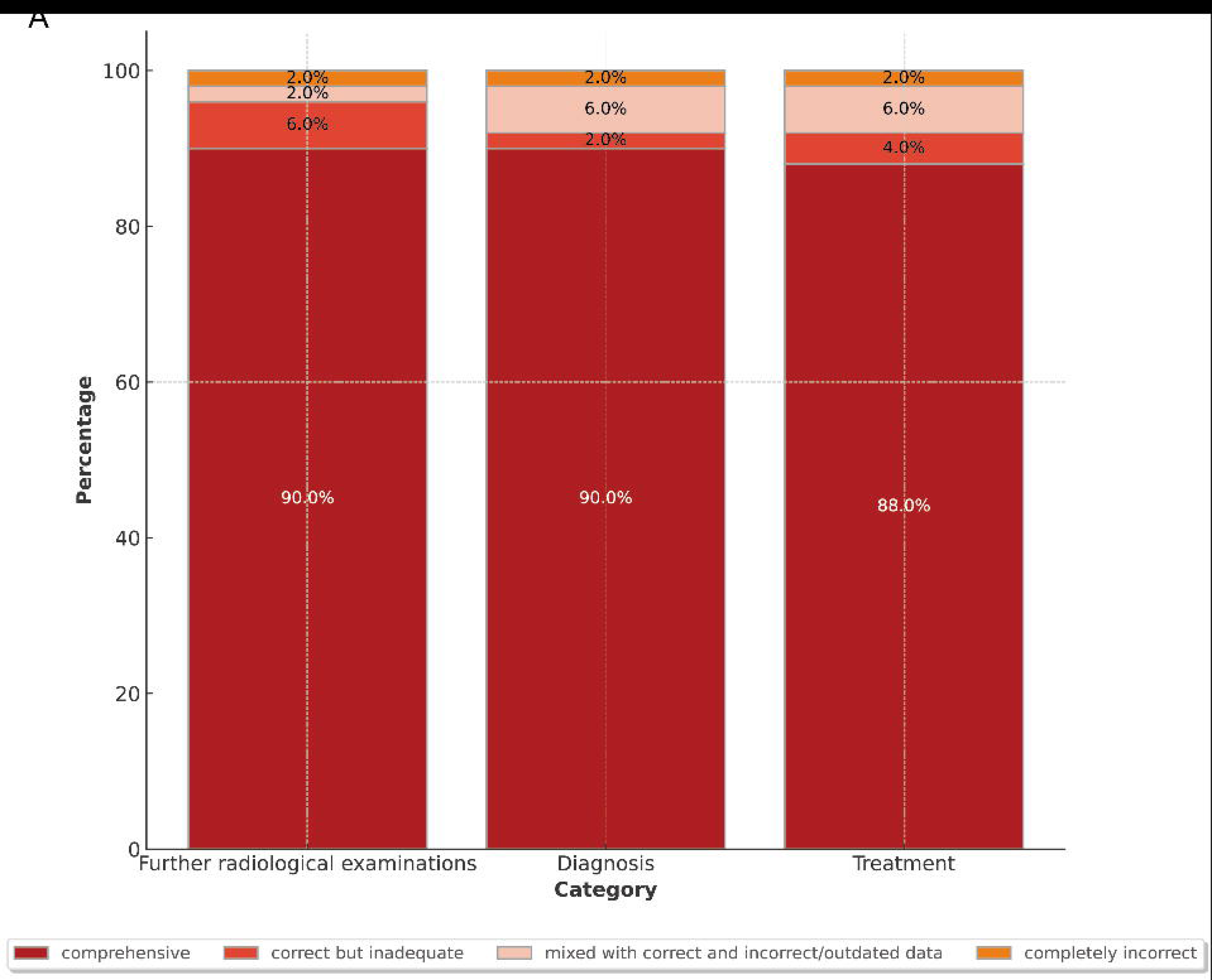
(A) The pie chart showed the accuracy rate of recommended level predicted by GPT-4. (B) Confusion matrix compared guideline recommended level with those predicted by GPT-4. (C) Bar chart showed Likert scale score distribution of accuracy. (D) Bar chart showed Likert scale score distribution of completeness.

**Table 1:**
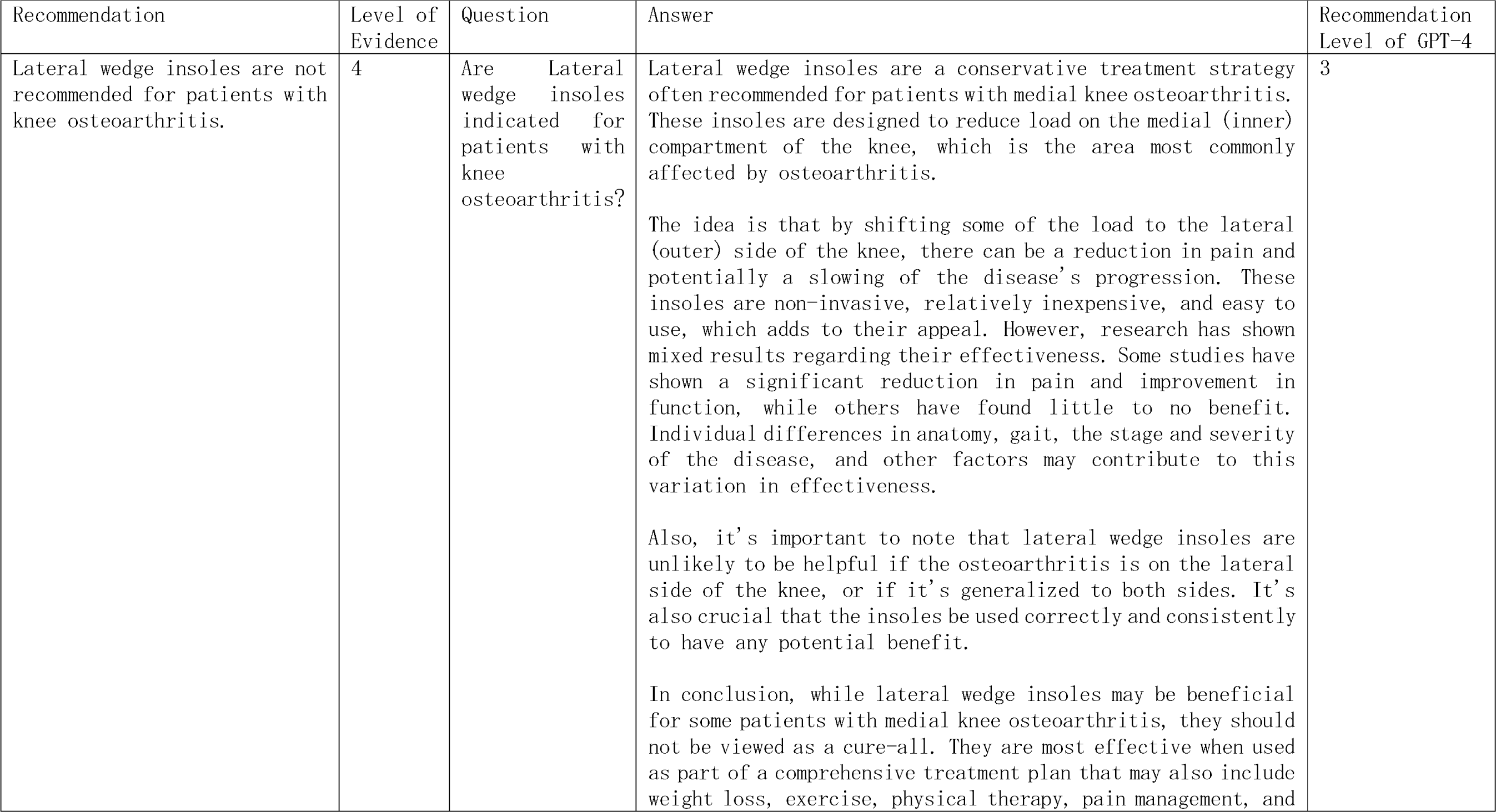
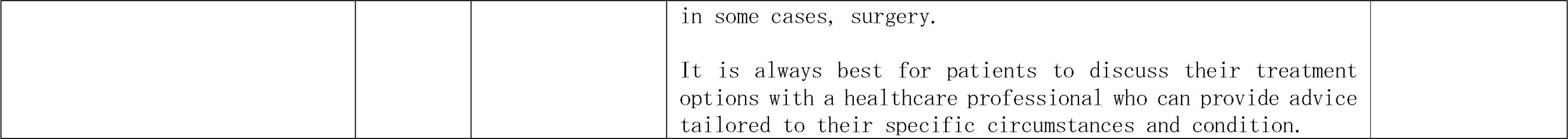
Examples of recommendation by AAOS, question, answer and recommendation level by GPT-4.

Figure 1C&D delineates the distribution of Likert scores for both accuracy and completeness. Out of the 28 responses generated by GPT4, the average score for precision was 4.3±1.6, and the average score for completeness stood at 2.8± 0.6(Table 4). The scores pertaining to accuracy and thoroughness did not exhibit significant variances across different levels of evidence or recommendation gradings.

### 1.2 Chinese guideline for diagnosis and treatment of osteoarthritis (2021 edition)

In the Chinese guidelines for OA, 15 key questions were proposed, with respect to which experts succinctly formulated 30 recommendations. In this study, these 15 questions were directly input into GPT-4 to explore the accuracy and completeness of its answers in relation to the 30 recommendations(Table 2 and Supplementary Table 2). Among the 30 recommendations, there were 11 rated as A-level, 11 as B-level, and eight as C-level. In terms of accuracy, the average scores of the three levels in GPT-4’s responses were 4.0±0.6, 4.5±0.6, and 4.5±0.7 respectively. In terms of completeness, the average scores of the three levels were 2.9±0.3, 2.3±0.9, and 2.1±1.0 respectively(Table 3). As shown in Figures 2A and 2B, most of the responses possess high accuracy and comprehensiveness, suggesting that GPT-4 provides comprehensive and precise answers to questions related to OA, reflecting a thorough understanding of OA.

**Figure 2:**
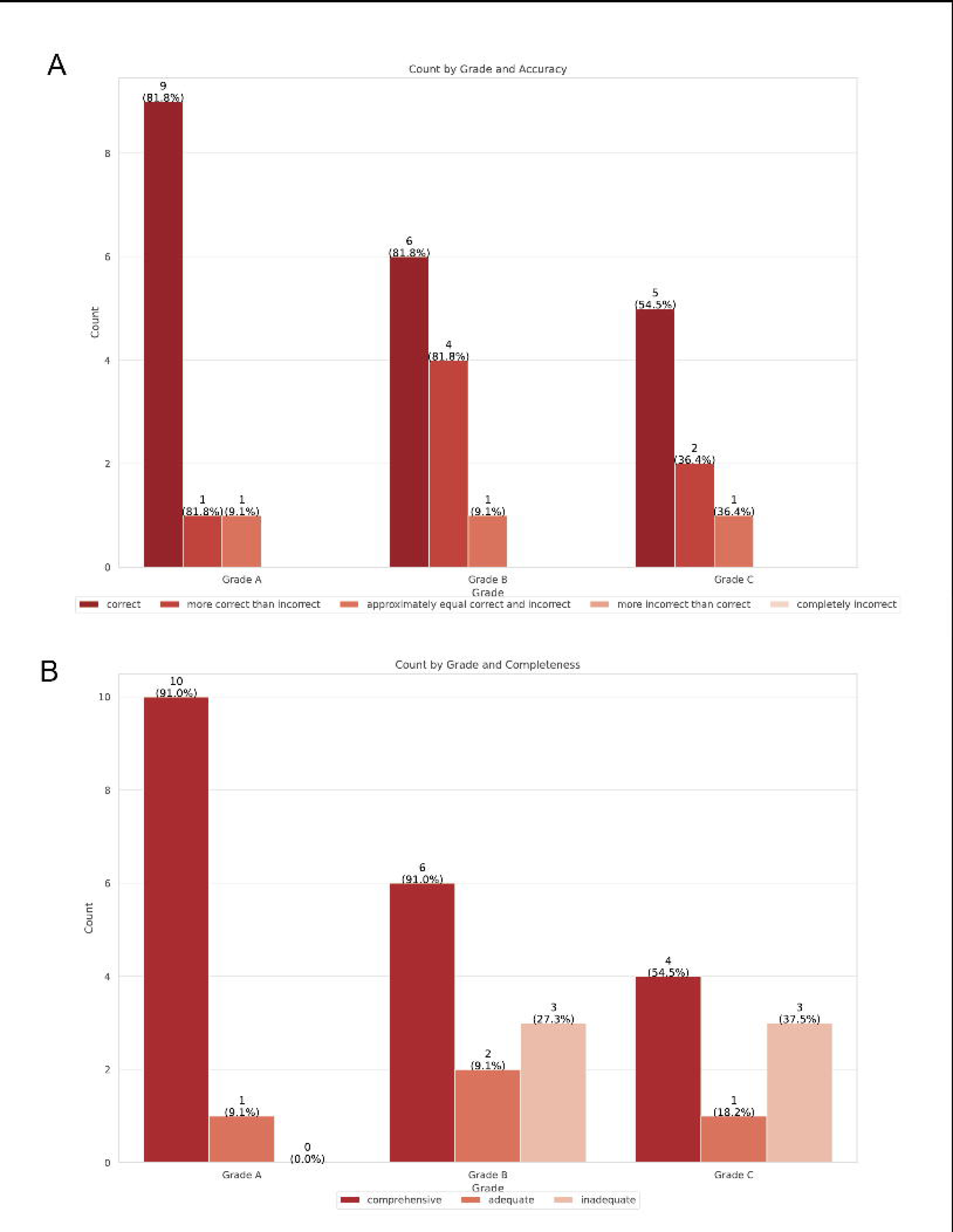
(A) The clustered bar graph illustrates the accuracy of GPT-4’s responses in proposing suggestions for GRADE A, B, and C levels.(B) The clustered bar graph illustrates the completeness of GPT-4’s responses in proposing suggestions for GRADE A, B, and C levels.

**Table 2:** Example of questions and recommendations from China guideline of OA, response from GPT-4.

| Question | Recommendation | GPT-4 response |
| --- | --- | --- |
| What imaging examinations should clinical physicians select for suspected osteoarthritis patients? | Recommendation 7:X-ray examination is the preferred choice for suspected osteoarthritis (OA) patients. If necessary, further examinations such as CT, MRI, and ultrasound can be performed to determine the location and extent of degeneration and facilitate differential diagnosis (strong recommendation, evidence level: B). | Imaging is an essential part of the diagnostic process for osteoarthritis (OA). The type of imaging depends on the suspected location of the OA, the severity of the patient's symptoms, and what information the doctor is trying to gather. Here are some common types of imaging used:<br>**\*\*X-ray\*\***: This is the most commonly used imaging technique for diagnosing OA. It can show bone damage, cartilage loss (seen as a narrowing of the space between the bones), bone spurs, and changes in the shape or alignment of the joints.<br>**\*\*MRI (Magnetic Resonance Imaging)\*\***: While not typically the first choice, an MRI can provide a more detailed image and show damage to other structures within the joint like ligaments and tendons, in addition to the bones and cartilage. MRI might be more helpful in the early stages of the disease when X-ray findings are not yet significant, or when more information is needed about the condition of the joint.<br>**\*\*Ultrasound\*\***: This is less commonly used for OA, but it can be helpful in evaluating soft tissue structures around the joint, such as bursae, and can also reveal fluid within the joint or bone spurs.<br>**\*\*CT scan (Computed Tomography)\*\***: While not commonly used for routine evaluation of OA, it can be helpful in some cases to assess the bony anatomy of the joint, particularly before joint replacement surgery.<br>In addition to imaging studies, the clinical diagnosis of OA often involves a medical history assessment, physical examination, and sometimes laboratory tests to help rule out other diseases. The choice of imaging will be guided by the physician based on the individual patient's circumstances. |

**Table 3:** GPT-4 accuracy and completeness against OA guideline from America and China.

| Osteoarthritis Guideline |  | Accuracy(5 points) | Completeness( 3 points) |
| --- | --- | --- | --- |
| AAOS |  | 4.3±1.6 | 2.8±0.6 |
| China Guideline | Grade A | 4.0±0.6 | 2.9±0.3 |
|  | Grade B | 4.5±0.6 | 2.3±0.9 |
|  | Grade C | 4.5±0.7 | 2.1±1.0 |

### 1.3 Case inquiry ability evaluation

In this study, we randomly selected 50 common orthopedic clinical cases, utilizing GPT-4 for case analysis to generate responses regarding further radiological examinations, diagnosis, and treatment(Table 4 and Supplementary Table 3). Across the three categories, GPT-4’s average scores were 3.78±0.52, 3.82±0.48, and 3.8±0.6, respectively. Figure 3A displays the quality of GPT-4’s responses, revealing a high level of performance across all categories, with over 88% of responses being comprehensive. GPT-4 only committed an error in the ’Further radiological examinations’ category in the case of peroneal nerve paralysis post-knee arthroplasty. Additionally, a diagnostic error occurred in the case of lumbar tuberculosis, which subsequently led to an incorrect treatment suggestion.

**Table 4:**
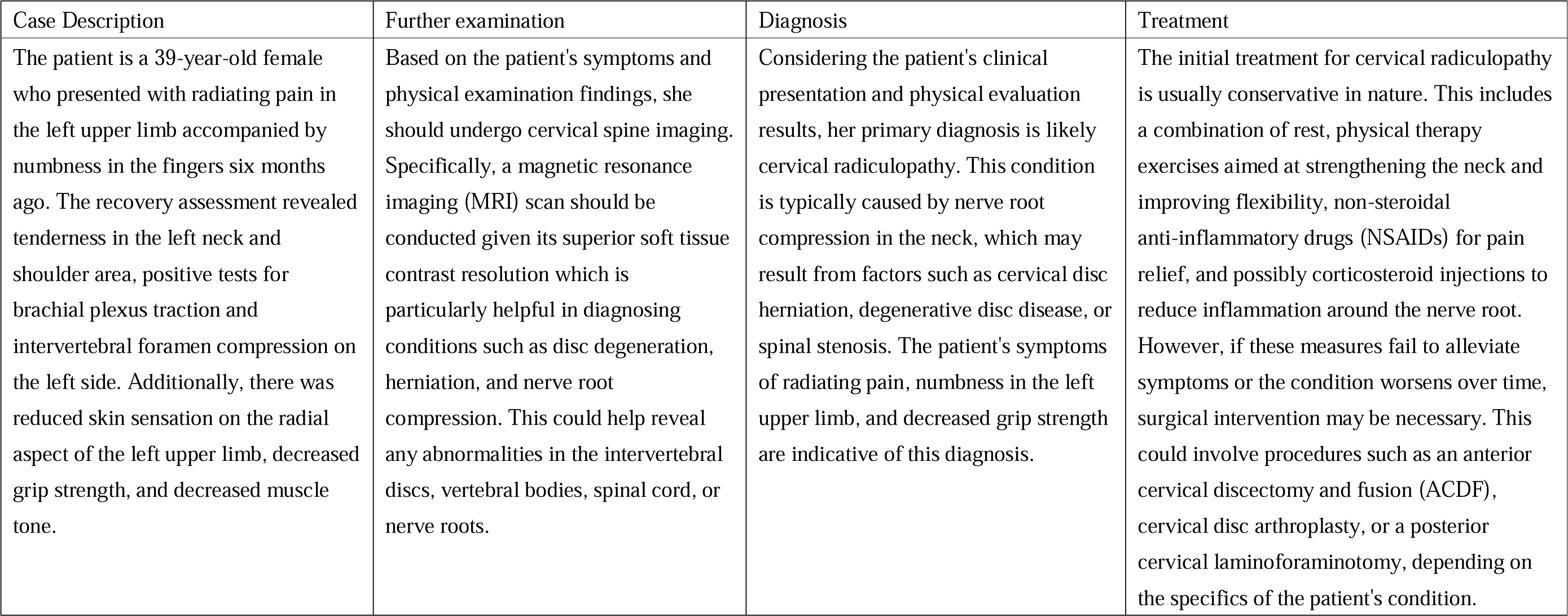
Example of orthopaedic cases and response by GPT-4.

| Case Description | Further examination | Diagnosis | Treatment |
| --- | --- | --- | --- |
| The patient is a 39-year-old female who presented with radiating pain in the left upper limb accompanied by numbness in the fingers six months ago. The recovery assessment revealed tenderness in the left neck and shoulder area, positive tests for brachial plexus traction and intervertebral foramen compression on the left side. Additionally, there was reduced skin sensation on the radial aspect of the left upper limb, decreased grip strength, and decreased muscle tone. | Based on the patient's symptoms and physical examination findings, she should undergo cervical spine imaging. Specifically, a magnetic resonance imaging (MRI) scan should be conducted given its superior soft tissue contrast resolution which is particularly helpful in diagnosing conditions such as disc degeneration, herniation, and nerve root compression. This could help reveal any abnormalities in the intervertebral discs, vertebral bodies, spinal cord, or nerve roots. | Considering the patient's clinical presentation and physical evaluation results, her primary diagnosis is likely cervical radiculopathy. This condition is typically caused by nerve root compression in the neck, which may result from factors such as cervical disc herniation, degenerative disc disease, or spinal stenosis. The patient's symptoms of radiating pain, numbness in the left upper limb, and decreased grip strength are indicative of this diagnosis. | The initial treatment for cervical radiculopathy is usually conservative in nature. This includes a combination of rest, physical therapy exercises aimed at strengthening the neck and improving flexibility, non-steroidal anti-inflammatory drugs (NSAIDs) for pain relief, and possibly corticosteroid injections to reduce inflammation around the nerve root. However, if these measures fail to alleviate symptoms or the condition worsens over time, surgical intervention may be necessary. This could involve procedures such as an anterior cervical discectomy and fusion (ACDF), cervical disc arthroplasty, or a posterior cervical laminoforaminotomy, depending on the specifics of the patient's condition. |

**Figure 3:**
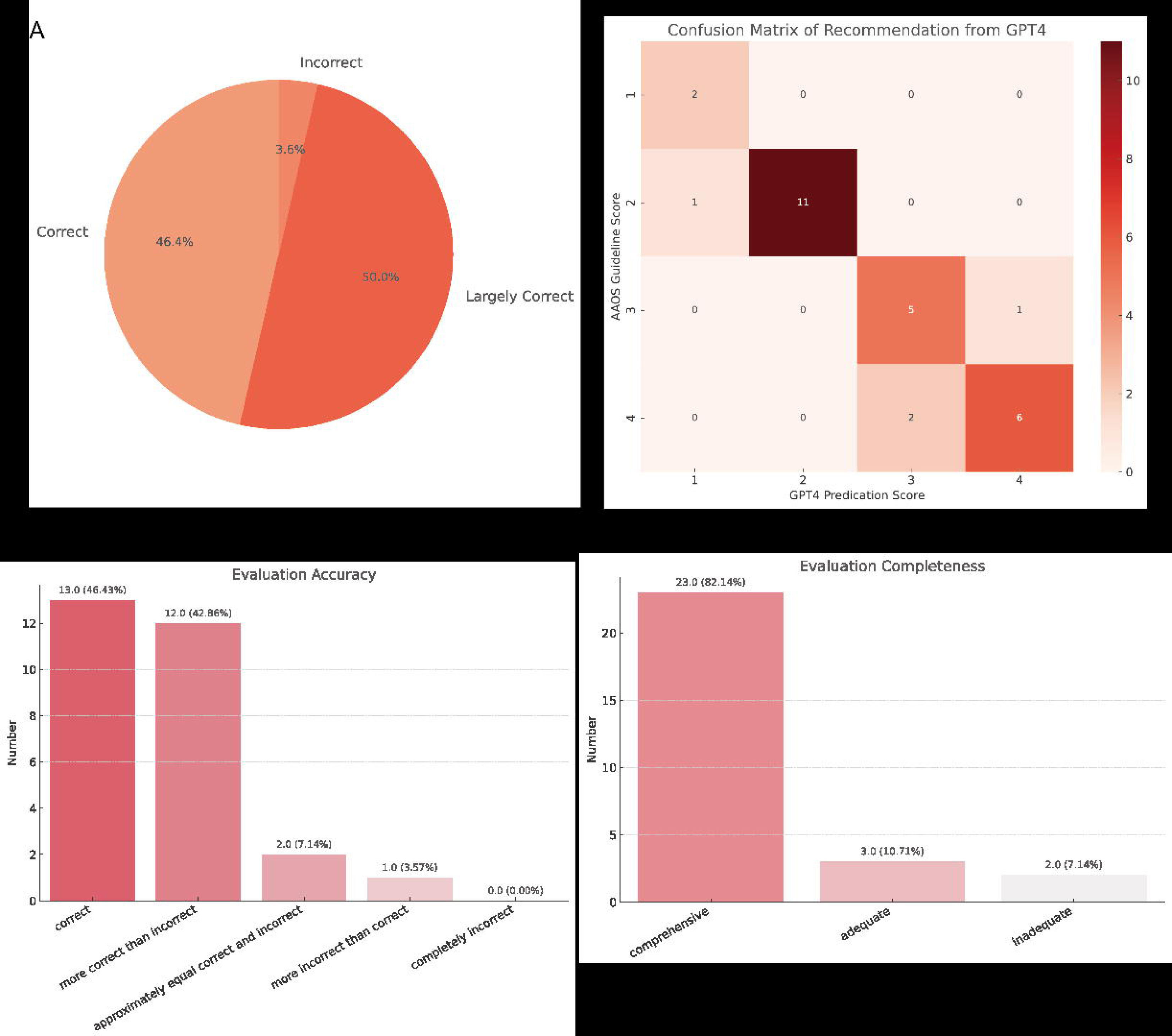
(A) The stacked bar chart shows the comprehensive level of GPT-4’s answers in the areas of further radiological examinations, primary diagnoses, and treatment.

## Discussion

The advent of AI, specifically GPT-4, has presented transformative potential for various fields, including medicine[23–25]. As an emerging innovation, GPT-4 necessitates thorough exploration and comprehensive validation prior to its incorporation into patient healthcare services. In this study, we sought to evaluate the efficacy of GPT-4 against the osteoarthritis treatment guidelines from both America and China and evaluate its ability of case inquiry.

The results of this study provide evidence of the potential utility and effectiveness of GPT-4 in the field of orthopedics, particularly with regards to OA. The impressive performance of GPT-4 in interpreting OA guidelines, responding to questions, and handling clinical cases demonstrates its potential as a supportive tool in orthopedic clinical practice. The case inquiry ability evaluation further highlights the potential of GPT-4 as a tool for clinical case analysis. While there were a few errors, the overall performance in suggesting further radiological examinations, providing diagnoses, and proposing treatment plans was highly commendable. It is noteworthy that even seasoned clinicians may make occasional errors, suggesting that GPT-4’s performance is comparable to human experts in certain respects.

GPT-4 indeed exhibits remarkable outcomes. For instance, it evinces a profound comprehension of the utility of traditional Chinese Medicine and herbal therapies in the investigation and management of osteoarthritis. In undertaking additional assessments for instances of post-joint replacement infection, GPT-4 explicitly articulates that C-reactive protein (CRP) and erythrocyte sedimentation rate (ESR) tests are required in conjunction with radiological examinations. Notably, through text-based case analysis alone, it possesses the capability to diagnose Felty’s syndrome accurately, a rare autoimmune disorder typically prevalent among individuals suffering from severe rheumatoid arthritis.

Other researchers from various medical fields have also explored the response capabilities of GPT-4, resulting in a myriad of perspectives. For example, Yuki et al evaluated GPT-4’s accuracy and completeness against the International Consensus Statement on Allergy and Rhinology: Rhinosinusitis[26]. However, only 54% of GPT-4’s responses achieved full marks in accuracy, and 71% received full marks in completeness. Yee et al used ChatGPT (GPT-3.5) to inquire about two diseases, cirrhosis and hepatocellular carcinom[27]. Both diseases achieved over 70% accuracy full marks and more than 40% completeness full marks. The authors believe that ChatGPT may serve as an adjunct informational tool for patients and physicians to improve outcomes.

In the field of orthopedics, although specific data is not yet available, there are already scholars who have made a certain degree of forecasts. For instance, GPT-4 can assist doctors in five areas within joint replacement: scientific research, disease diagnosis, treatment options, preoperative planning, intraoperative support, and postoperative rehabilitation[28]. In sports medicine, GPT-4 can contribute to diagnostic imaging, exercise prescription, medical supervision, surgical treatment, sports nutrition, and scientific research[29]. The author believes that while GPT-4 will not replace doctors, it could become an indispensable scientific assistant for sports doctors in the future.

However, while these findings are promising, it’s important to approach the integration of AI tools like GPT-4 in healthcare with caution. A few errors identified in the case analysis suggest that the tool is not infallible and should not be relied upon blindly. Human oversight and supervision remain essential, particularly in complex and nuanced clinical scenarios. It is also important to consider that the tool’s performance could be influenced by the quality and specificity of the input data provided. Therefore, continued research and monitoring of GPT-4’s performance in different clinical situations and contexts is necessary.

In conclusion, this study provides promising insights into the application of AI, particularly GPT-4, in the field of orthopedics. As technology continues to advance, it could potentially revolutionize healthcare delivery, making it more efficient and precise. However, it is crucial that we carefully evaluate and monitor these tools to ensure that they are used responsibly and effectively.

## Data Availability

All data produced in the present study are available upon reasonable request to the authors

