## supplemental Table 1 for "Assessing the performance of GPT-4 in the filed of osteoarthritis and orthopaedic case consultation"

Supplementary Table 1: Recommendation by AAOS, question, answer and recommendation level by GPT-4.

|  | Recommendation | Level of Evidence | Question | Answer | Recommendation Level of GPT-4 |
| --- | --- | --- | --- | --- | --- |
| 1 | Lateral wedge insoles are not recommended for patients with knee osteoarthritis. | 4 | Are Lateral wedge insoles indicated for patients with knee osteoarthritis? | Lateral wedge insoles are a conservative treatment strategy often recommended for patients with medial knee osteoarthritis. These insoles are designed to reduce load on the medial (inner) compartment of the knee, which is the area most commonly affected by osteoarthritis.  The idea is that by shifting some of the load to the lateral (outer) side of the knee, there can be a reduction in pain and potentially a slowing of the disease's progression. These insoles are non-invasive, relatively inexpensive, and easy to use, which adds to their appeal. However, research has shown mixed results regarding their effectiveness. Some studies have shown a significant reduction in pain and improvement in function, while others have found little to no benefit. Individual differences in anatomy, gait, the stage and severity of the disease, and other factors may contribute to this variation in effectiveness.  Also, it's important to note that lateral wedge insoles are unlikely to be helpful if the osteoarthritis is on the lateral side of the knee, or if it's generalized to both sides. It's also crucial that the insoles be used correctly and consistently to have any potential benefit.  In conclusion, while lateral wedge insoles may be beneficial for some patients with medial knee osteoarthritis, they should not be viewed as a cure-all. They are most effective when used as part of a comprehensive treatment plan that may also include weight loss, exercise, physical therapy, pain management, and in some cases, surgery.  It is always best for patients to discuss their treatment options with a healthcare professional who can provide advice tailored to their specific circumstances and condition. | 3 |
| 2 | Canes could be used to improve pain and function in patients with knee osteoarthritis. | 3 | Are canes recommended for improving function and quality of osteoarthritis patient? | Yes, canes can be a useful tool to manage knee or hip osteoarthritis. They can help by reducing the load placed on the affected joint, thereby reducing pain and potentially slowing the disease's progression.  By providing additional support and balance, a cane can improve mobility and safety for patients with osteoarthritis. This can lead to improved function, as patients may be able to engage in activities that were previously difficult due to pain or instability.  Additionally, the use of a cane can help improve a patient's quality of life. By reducing pain and increasing mobility, a cane can allow patients to maintain their independence and continue to participate in activities they enjoy.  It's important, however, that the cane is used correctly. Incorrect use can lead to other problems, such as shoulder or arm pain. Generally, the cane should be held in the hand opposite to the affected leg. This allows the cane to bear weight when the affected leg is in the swing phase of gait. Also, the height of the cane needs to be appropriate for the individual, typically set at the level of the wrist when the arm is hanging naturally at the side.  As with all treatments for osteoarthritis, the use of a cane should be discussed with a healthcare professional to ensure it is appropriate and beneficial for the individual patient. Canes are typically part of a larger treatment plan that may include medication, physical therapy, exercise, weight management, and possibly surgery. | 3 |
| 3 | Brace treatment could be used to improve function, pain, and quality of life in patients with knee osteoarthritis | 3 | Are brace indicated for knee osteoarthritis to improve function, pain and quality of life in patient | Yes, knee braces are often used as a non-invasive, non-pharmacological treatment option for knee osteoarthritis. They are designed to improve function, reduce pain, and potentially slow the progression of the disease by altering force distributions across the knee joint.  There are different types of knee braces that can be used depending on the specific needs of the patient:  1. Unloader (or off-loader) Braces: These are designed to shift weight away from the affected part of the knee. They are often used in patients with unicompartmental knee osteoarthritis (most commonly medial compartment). By off-loading the weight to the healthier compartment, they can help reduce pain and improve function.  2. Support Braces: These are more general braces designed to provide overall support to the knee, reduce swelling, and ease pain. They can be helpful in cases where the osteoarthritis is more diffuse and not limited to one compartment.  The effectiveness of knee braces can vary from person to person. Some people find significant relief and improved function, while others find little benefit. Factors influencing their effectiveness can include the severity and location of the arthritis, the design and fit of the brace, and patient adherence to the treatment plan.  It's also worth noting that while braces can provide symptomatic relief, they do not cure the underlying disease. Other treatments such as pain medication, physical therapy, lifestyle modifications (e.g., weight loss, exercise), and potentially surgery may also be necessary.  As always, patients should consult with their healthcare provider to determine the most appropriate treatment plan for their specific condition. It's important for knee braces to be properly fitted by a professional to ensure maximum effectiveness and comfort. | 3 |
| 4 | The following supplements may be helpful in reducing pain and improving function for patients with mild to moderate knee osteoarthritis; however, the evidence is inconsistent/limited and additional research clarifying the efficacy of each supplement is needed. • Turmeric • Ginger extract • Glucosamine • Chondroitin • Vitamin D | 2 | Are supplements such as turmeric, ginger extract, recommended for osteoarthritis patients for reducing pain and improving function | Dietary supplements are frequently used as an adjunctive treatment for osteoarthritis, including turmeric (curcumin) and ginger extract. Both have been studied for their anti-inflammatory properties and potential benefits in managing osteoarthritis symptoms.  Turmeric contains the active compound curcumin, which is known for its potent anti-inflammatory and antioxidant properties. Some studies have suggested that curcumin can help reduce pain and improve function in people with osteoarthritis, though the evidence is not as robust or consistent as that for traditional treatments like NSAIDs.  Ginger extract also has anti-inflammatory properties and has been used traditionally for various ailments, including joint pain. Some research suggests that ginger may be beneficial in managing osteoarthritis symptoms. However, like turmeric, the evidence is not as consistent or robust as for some other treatments.  It's important to note that while these supplements might help manage symptoms of osteoarthritis, they are not a cure for the underlying disease. They should be used as part of a comprehensive treatment plan that may also include physical therapy, exercise, weight management, and possibly medication or surgery.  Patients should also be aware that dietary supplements are not regulated in the same way as prescription medications. The quality, purity, and concentration of active ingredients can vary between brands.  Furthermore, while generally considered safe, these supplements can have side effects and interact with other medications. For example, both turmeric and ginger can thin the blood and may increase the risk of bleeding, especially if taken with other blood-thinning medications.  Therefore, it's essential to talk with a healthcare provider before starting any new supplement regimen. They can provide advice on safe and effective dosages and monitor for potential side effects or drug interactions. | 1 or 2 |
| 5 | Topical NSAIDs should be used to improve function and quality of life for treatment of osteoarthritis of the knee, when not contraindicated. | 4 | Are topical NSAIDs indicated for improving function and quality of osteoarthritis patient? | Yes, topical nonsteroidal anti-inflammatory drugs (NSAIDs) are often used as part of the treatment plan for osteoarthritis, particularly for patients with osteoarthritis of the knees or hands.  Topical NSAIDs work by reducing the production of prostaglandins, substances in the body that cause inflammation and increase pain sensitivity. By applying the NSAID directly to the area of pain, it is thought to concentrate the medicine at the site of pain and inflammation while reducing the amount absorbed into the bloodstream and thus potentially reducing systemic side effects.  Several studies have found topical NSAIDs to be effective in relieving osteoarthritis pain and improving function. In fact, guidelines from professional organizations such as the American College of Rheumatology and the Arthritis Foundation recommend topical NSAIDs as a first-line treatment for knee or hand osteoarthritis, particularly for patients at increased risk of gastrointestinal side effects from oral NSAIDs.  Examples of topical NSAIDs include diclofenac gel (Voltaren) and patches, and they are available both over the counter and by prescription, depending on the country and specific product.  As with all medications, there can be side effects. Topical NSAIDs can cause local skin irritation, and while less common than with oral NSAIDs, systemic effects such as gastrointestinal upset or cardiovascular effects can occur, particularly with prolonged use or in people with underlying risk factors.  It's always important for patients to discuss their treatment options with a healthcare provider who can provide advice based on their specific circumstances and overall health status. | 4 |
| 6 | Supervised exercise, unsupervised exercise, and/or aquatic exercise are recommended over no exercise to improve pain and function for treatment of knee osteoarthritis. | 4 | Are different type of exercise recommended instead of no exercise for improving function and quality of osteoarthritis patient? | Yes, exercise is a key component of managing osteoarthritis and is widely recommended to improve function and quality of life. Exercise can help to reduce pain, improve mobility, increase muscle strength, enhance balance and coordination, and promote overall health and wellbeing. It can also assist in weight management, which is particularly important in osteoarthritis as excess weight can put additional strain on weight-bearing joints.  There are several types of exercise that can be beneficial for patients with osteoarthritis:  1. Aerobic Exercise: This includes activities such as walking, cycling, swimming, or using an elliptical machine. Aerobic exercise can improve cardiovascular fitness, help control weight, and improve overall function.  2. Strength Training: Resistance exercises can strengthen the muscles that support affected joints, which can reduce pain and improve function. This can be performed using weights, resistance bands, or body weight exercises.  3. Flexibility Exercises: Stretching exercises can help to maintain or improve the flexibility of the joints and surrounding muscles, which can assist in maintaining function and mobility.  4. Balance and Proprioception Exercises: These exercises can enhance balance and joint awareness, which can reduce the risk of falls and improve function.  5. Aquatic Exercise: For individuals with significant pain or difficulty with land-based exercise, aquatic exercise can be a good alternative. The buoyancy of the water reduces stress on the joints while providing resistance for strength training.  It's important to note that the type, intensity, and duration of exercise should be individualized based on the patient's overall health, fitness level, and severity of osteoarthritis. It's also crucial to start slowly and gradually increase intensity and duration to reduce the risk of injury or increased pain.  Patients should consult with their healthcare provider or a physical therapist to develop an exercise plan that suits their individual needs and capabilities. Also, it's key to remember that exercise is just one part of a comprehensive treatment plan for osteoarthritis, which may also include pain management, use of assistive devices, and possibly surgery. | 4 |
| 7 | Neuromuscular training (i.e. balance, agility, coordination) programs in combination with traditional exercise could be used to improve performance based function and walking speed for treatment of knee osteoarthritis. | 3 | Are neuromuscular training in combination with traditional exercise indicated for improving function and quality of osteoarthritis patient? | Yes, neuromuscular training can be a beneficial addition to a traditional exercise program for patients with osteoarthritis, particularly those with knee and hip osteoarthritis. Neuromuscular training focuses on improving muscle control, coordination, balance, proprioception (the ability to sense the position and movement of the joints), and the ability to react to changing conditions or uneven surfaces, all of which can enhance function and reduce the risk of falls and further injury.  Research has shown that neuromuscular training can help to reduce pain and improve function in people with osteoarthritis. It can help to improve the alignment and stability of the affected joint, potentially reducing the abnormal loading that contributes to the progression of osteoarthritis.  Neuromuscular training often includes balance exercises, agility drills, proprioceptive training (for example, exercises on unstable surfaces like a balance board), and specific exercises to improve the control and coordination of the muscles that support the affected joint.  This type of training should be individualized to the patient's specific needs, capabilities, and goals, and ideally should be guided by a physical therapist or other healthcare professional with expertise in this area.  As with all exercise programs for osteoarthritis, it's important for the program to be part of a comprehensive treatment plan that may also include weight management, pain management, and possibly other interventions such as assistive devices or surgery. It's also important for patients to consult with their healthcare provider before starting any new exercise regimen.  In conclusion, while more research is needed to fully understand the optimal design and benefits of neuromuscular training programs for people with osteoarthritis, current evidence suggests that it can be a valuable component of an overall management strategy for this condition. | 3 |
| 8 | Self- management programs are recommended to improve pain and function for patients with knee osteoarthritis. | 4 | Are self-management programs recommended for improving function and quality of osteoarthritis patient? | Yes, self-management programs are highly recommended for patients with osteoarthritis. These programs empower patients to take an active role in managing their disease, which can lead to improved function, pain control, and overall quality of life.  Self-management programs typically include education about the disease, techniques for joint protection, pain management strategies, physical activity recommendations, nutritional advice, stress management techniques, and communication skills to effectively talk with healthcare providers.  Several studies and reviews have demonstrated the benefits of self-management programs for osteoarthritis patients. These benefits include decreased pain, improved physical function, increased knowledge about osteoarthritis, improved self-efficacy (confidence in one's ability to manage the disease), and overall improved quality of life.  However, for self-management programs to be successful, they must be individualized to each patient's needs, abilities, and resources. They should also be delivered in a supportive, patient-centered manner that empowers the patient to take an active role in their care.  Patients should work with their healthcare providers to develop a self-management plan that suits their specific circumstances. This might involve a team of healthcare providers, including doctors, nurses, physical therapists, dietitians, and possibly others.  Given the strong evidence supporting the benefits of self-management programs, if we were to grade their recommendation level with 4 being the highest, it would likely be close to a 4, although the exact grading could depend on the specific details of the grading system. It's also important to note that self-management programs should be part of a comprehensive treatment approach that may also include medication, physical therapy, assistive devices, and possibly surgery. | 4 |
| 9 | Patient education programs are recommended to improve pain in patients with knee osteoarthritis. | 4 | Are patient education programs recommended for improving function and quality of osteoarthritis patient? | Yes, patient education programs are a key component of the management of osteoarthritis and are highly recommended. These programs aim to increase patients' knowledge about the disease, help them understand their treatment options, and empower them to take an active role in their own care.  Patient education programs can cover a range of topics, including:  Understanding the nature of osteoarthritis  Learning about the various treatment options and their risks and benefits  Managing pain and other symptoms  Implementing lifestyle changes, such as diet and exercise modifications  Using assistive devices, if needed  Techniques for protecting joints and conserving energy  Developing coping strategies for living with a chronic disease  Several studies have shown that patient education programs can improve outcomes for people with osteoarthritis. They can help to reduce pain, improve physical function, increase adherence to treatment plans, and enhance overall quality of life. | 4 |
| 10 | Sustained weight loss is recommended to improve pain and function in overweight and obese patients with knee osteoarthritis. | 3 | Are sustained weight loss recommended for improving function and quality of osteoarthritis patient? | Yes, sustained weight loss is strongly recommended for patients with osteoarthritis who are overweight or obese.  Excess weight places additional strain on weight-bearing joints such as the knees and hips, which can exacerbate the symptoms of osteoarthritis and potentially accelerate the progression of the disease.  Research has shown that weight loss can reduce pain, improve physical function, and enhance quality of life in people with knee osteoarthritis. The Arthritis Foundation suggests that for every pound of weight loss, there's a four-pound reduction in the load exerted on the knee for each step taken during daily activities. Therefore, even modest weight loss can have a significant impact on reducing joint stress.  Furthermore, weight loss can improve general health and reduce the risk of other health conditions that are common in people with osteoarthritis, such as cardiovascular disease and diabetes.  It's important to note that weight loss should be achieved and maintained through a combination of a healthy, balanced diet and regular physical activity. Crash diets or excessive exercise can be harmful, particularly for individuals with osteoarthritis. A healthcare provider or a dietitian can provide advice on safe and effective ways to lose weight and maintain weight loss. | 4 |
| 11 | Manual therapy in addition to an exercise program may be used to improve pain and function in patients with knee osteoarthritis. | 2 | Are manual therapy in addition to an exercise program indicated improving function and quality of osteoarthritis patient? | Manual therapy, when used in conjunction with an exercise program, can be a beneficial part of the treatment plan for some patients with osteoarthritis, particularly those with hip and knee osteoarthritis.  Manual therapy is a physical treatment used by physiotherapists to treat musculoskeletal pain and disability, and it involves hands-on techniques to manipulate joints, mobilize soft tissues, and reduce pain. These techniques can include massage, stretching, resistance movements, and joint manipulations or mobilizations.  For osteoarthritis patients, manual therapy can help to reduce pain, improve range of motion, increase flexibility, and improve function. When used in combination with a structured exercise program, it can help to enhance the benefits of the exercises and may lead to better outcomes in terms of pain and function.  However, it's important to note that the evidence supporting the use of manual therapy for osteoarthritis is somewhat mixed. Some studies have found it to be beneficial, while others have found little to no effect. The success of the therapy may depend on a variety of factors, including the specific techniques used, the severity of the osteoarthritis, the joints involved, and the overall health and fitness of the patient. | 2 or 3 |
| 12 | Massage may be used in addition to usual care to improve pain and function in patients with knee osteoarthritis. | 2 | Are massage indicated to improve function and quality of osteoarthritis patient in addition to usual care | Massage therapy, as a complementary approach to usual care, can potentially provide some benefits to patients with osteoarthritis. Massage can help to relax muscles, improve circulation, and reduce stress, which can contribute to pain relief and improved function.  Several studies have shown that massage therapy can reduce pain and improve function in people with knee and hip osteoarthritis. A systematic review and meta-analysis published in Pain Medicine in 2020 found that massage therapy can provide short-term relief of knee osteoarthritis pain.  However, massage therapy should not replace conventional treatments for osteoarthritis but rather serve as an additional tool for managing symptoms and improving quality of life. Moreover, it's important to ensure that massage therapy is performed by a qualified professional who understands osteoarthritis, as inappropriate techniques can potentially cause harm.  The effectiveness of massage therapy can vary depending on the specific techniques used, the duration and frequency of the sessions, and the individual patient's response to treatment. Therefore, patient preference and individual circumstances should be taken into account when considering the addition of massage therapy to an osteoarthritis treatment plan. | 2 or 3 |
| 13 | FDA-approved laser treatment may be used to improve pain and function in patients with knee osteoarthritis. | 2 | Are FDA-approved laser treatment indicated for improving function and quality of osteoarthritis patient? | Low-level laser therapy (LLLT), also known as cold laser therapy, has been studied for the treatment of pain and inflammation related to osteoarthritis. The mechanism behind this treatment is thought to be related to its ability to influence cellular function, promote tissue repair, and decrease inflammation and pain.  As of my knowledge cutoff in September 2021, some clinical studies have shown that low-level laser therapy can reduce pain and improve function in people with knee osteoarthritis. However, the overall evidence is somewhat mixed, with some studies showing benefits and others showing no significant effect.  One of the challenges in interpreting the evidence is that studies have used different types of lasers, different treatment protocols (such as the power of the laser, the duration of treatment, and the number of treatment sessions), and different outcome measures.  The Food and Drug Administration (FDA) in the United States has cleared some devices for the use of low-level laser therapy for the temporary relief of minor muscle and joint aches, pains, and stiffness, and for the temporary increase in local blood circulation. These clearances, however, do not indicate an endorsement of effectiveness for specific conditions like osteoarthritis. | 2 |
| 14 | Acupuncture may improve pain and function in patients with knee osteoarthritis. | 2 | Are acupuncture indicated for improving function and quality of osteoarthritis patient? | Acupuncture, a traditional Chinese medicine technique that involves inserting thin needles into specific points on the body, has been studied for a variety of health conditions, including osteoarthritis.  As of my knowledge cutoff in September 2021, the research on acupuncture for osteoarthritis has provided mixed results. Some studies suggest it may provide some relief from osteoarthritis pain and improve function, particularly in the knee. However, other studies have not found a significant benefit, and some research suggests that the perceived benefits may be largely due to a placebo effect.  The American College of Rheumatology and Arthritis Foundation (ACR/AF) guidelines from 2019 conditionally recommend acupuncture for the management of osteoarthritis of the hand, hip, and knee. This suggests that while the evidence may not be as strong or consistent as for some other interventions, it might still be a reasonable option to consider for some patients, particularly those who have not responded to or cannot use standard treatments.  The World Health Organization (WHO) also recognizes acupuncture as a treatment for a variety of conditions, including symptomatic relief of osteoarthritis. | 2 or 3 |
| 15 | Modalities that may be used to improve pain and/or function in patients with knee osteoarthritis include: a. Transcutaneous Electrical Nerve Stimulation (pain) | 2 | Are Transcutaneous Electrical Nerve Stimulation recommended for improving function and quality of osteoarthritis patient? | Transcutaneous Electrical Nerve Stimulation (TENS) is a non-invasive method of pain relief commonly used in physical therapy. It involves the use of a device that sends small electrical currents to targeted body parts, which is believed to interrupt the nerve pathways that are sending pain signals to the brain.  As of my knowledge cutoff in September 2021, the evidence regarding the effectiveness of TENS for osteoarthritis is somewhat mixed. Some studies have found that TENS can provide short-term relief from osteoarthritis pain, but others have found no significant benefit. The quality of the available research is also variable, and there's a need for more high-quality, randomized controlled trials to definitively establish the effectiveness of TENS for osteoarthritis.  The American College of Rheumatology (ACR) and the Arthritis Foundation (AF) conditionally recommend TENS for the initial management of knee osteoarthritis. This suggests that while the evidence may not be as strong or consistent as for some other interventions, TENS might be a reasonable option to consider for some patients, particularly those who have not responded to or cannot use standard treatments. | 2 or 3 |
| 16 | Modalities that may be used to improve pain and/or function in patients with knee osteoarthritis include: a. Percutaneous Electrical Nerve Stimulation (pain and function) b. Pulsed Electromagnetic Field Therapy (pain) | 2 | Are Percutaneous Electrical Nerve Stimulation (pain and function) or Pulsed Electromagnetic Field Therapy (pain) indicated for improving function and quality of osteoarthritis patient? | Percutaneous Electrical Nerve Stimulation (PENS) and Pulsed Electromagnetic Field Therapy (PEMF) are two modalities that have been studied for managing pain and improving function in patients with osteoarthritis.  1. Percutaneous Electrical Nerve Stimulation (PENS): This is a technique that involves the insertion of thin acupuncture-like needles into the skin, which are then electrically stimulated. As of my knowledge cutoff in September 2021, there is limited evidence on the effectiveness of PENS for osteoarthritis. Some small studies suggest it might provide pain relief, but the overall quality and quantity of research is limited. More high-quality, large-scale studies are needed to confirm these results and to understand the potential benefits and risks of PENS for osteoarthritis.  2. Pulsed Electromagnetic Field Therapy (PEMF): This involves using a device that generates electromagnetic fields to promote healing and reduce inflammation and pain. Several studies have suggested that PEMF might be beneficial for reducing pain in people with knee osteoarthritis. A systematic review published in Physical Therapy in 2016 found that PEMF could reduce pain and improve physical function in the short term for individuals with knee osteoarthritis.  However, the evidence base for both of these modalities is not as strong or extensive as it is for some other interventions, like exercise, weight loss for overweight/obese individuals, and self-management programs. | 2 |
| 17 | Extracorporeal shockwave therapy may be used to improve pain and function for treatment of osteoarthritis of the knee. | 2 | Are Extracorporeal shockwave therapy indicated for improving function and quality of osteoarthritis patient? | Extracorporeal shockwave therapy (ESWT) is a non-invasive treatment that involves the delivery of shock waves to injured parts of the body to induce healing. It's often used in physical therapy and orthopedics for conditions like plantar fasciitis, tendinitis, and other musculoskeletal disorders.  As of my knowledge cutoff in September 2021, the research on ESWT for osteoarthritis is somewhat limited and has yielded mixed results. Some studies have found that ESWT can provide short-term relief from osteoarthritis pain and improve function, particularly in the knee. However, other studies have found no significant benefit, and the overall quality of the evidence is variable.  A systematic review and meta-analysis published in Archives of Physical Medicine and Rehabilitation in 2017 found that ESWT may improve pain, stiffness, and physical function in patients with knee osteoarthritis in the short term. However, the authors noted the need for further high-quality research to confirm these findings and to assess the long-term effects of ESWT. | 2 |
| 18 | Oral NSAIDs are recommended to improve pain and function in the treatment of knee osteoarthritis when not contraindicated. | 4 | Are Oral NSAIDs recommended for improving function and quality of osteoarthritis patient? | Oral non-steroidal anti-inflammatory drugs (NSAIDs), such as ibuprofen and naproxen, are commonly used to help manage symptoms of osteoarthritis, including pain and inflammation. They work by reducing the production of chemicals in the body that cause inflammation and pain.  As of my knowledge cutoff in September 2021, the American College of Rheumatology (ACR) and Arthritis Foundation (AF) guidelines recommend the use of oral NSAIDs for the management of osteoarthritis of the hand, hip, and knee. They can help relieve pain and improve function in individuals with osteoarthritis.  However, while these drugs can be effective, they also come with potential side effects and risks. These include gastrointestinal issues, such as stomach ulcers and bleeding, cardiovascular problems, and kidney damage with long-term use. Therefore, their use should be monitored by a healthcare provider, and they are often recommended for use at the lowest effective dose and for the shortest possible duration. | 4 |
| 19 | Oral acetaminophen is recommended to improve pain and function in the treatment of knee osteoarthritis when not contraindicated. | 4 | Are Oral acetaminophen recommended for improving function and quality of osteoarthritis patient? | Acetaminophen, also known as paracetamol in some countries and often sold under the brand name Tylenol, is a commonly used over-the-counter medication for relieving pain and reducing fever.  For osteoarthritis, acetaminophen has traditionally been recommended as a first-line treatment due to its safety profile when used at recommended dosages. It's generally well-tolerated and has fewer gastrointestinal side effects compared to non-steroidal anti-inflammatory drugs (NSAIDs).  However, while acetaminophen can help with mild to moderate osteoarthritis pain, it does not have anti-inflammatory properties, and thus may not be as effective as NSAIDs in managing osteoarthritis symptoms, particularly in more severe cases.  The American College of Rheumatology (ACR) and the Arthritis Foundation (AF) in their 2019 guidelines conditionally recommend against the use of acetaminophen for the management of osteoarthritis of the hand, hip, and knee. This decision was based on new evidence suggesting that its effectiveness for osteoarthritis pain may be less than previously thought.  Nonetheless, the decision to use acetaminophen should be individualized based on the patient's specific symptoms, overall health status, risk of side effects, and personal preferences. Some patients may still find acetaminophen to be a helpful part of their pain management strategy. | 1 or 2 |
| 20 | Oral narcotics, including tramadol, result in a significant increase of adverse events and are not effective at improving pain or function for treatment of osteoarthritis of the knee. | 4 | Are Oral narcotics, including tramadol recommended for improving function and quality of osteoarthritis patient? | Opioid analgesics, including tramadol and other narcotics, have been used in the treatment of osteoarthritis (OA) for severe pain that is unresponsive to other treatments. However, the use of opioids for chronic non-cancer pain, including OA, has become increasingly controversial due to concerns about side effects, addiction risk, and effectiveness.  As of my knowledge cutoff in September 2021, evidence suggests that while opioids may provide some pain relief for OA in the short term, their long-term efficacy is uncertain. Additionally, opioids are associated with a number of potential side effects, including drowsiness, nausea, constipation, dependency, and overdose. Over time, tolerance to these medications can develop, leading to the need for higher doses to achieve the same pain relief and increasing the risk of side effects.  The American College of Rheumatology (ACR) and the Arthritis Foundation (AF) in their 2019 guidelines strongly recommend against the use of opioids (excluding tramadol) for the management of osteoarthritis of the hand, hip, and knee due to the associated risks. The use of tramadol is conditionally recommended for patients who have not responded to other treatments.  Overall, while narcotics like tramadol may play a role in managing severe OA pain that's unresponsive to other treatments, they should be used with caution and typically as a last resort due to the significant potential for adverse events and limited evidence of long-term effectiveness. | 4 |
| 21 | Hyaluronic acid intra-articular injection(s) is not recommended for routine use in the treatment of symptomatic osteoarthritis of the knee. | 3 | Are Hyaluronic acid intra-articular injection(s) indicated for improving function and quality of osteoarthritis patient? | Hyaluronic acid (HA) intra-articular injections, also known as viscosupplementation, have been used as a treatment for osteoarthritis, especially for the knee. These injections are designed to supplement the viscosity of the synovial fluid in the joint, which can be decreased in osteoarthritis.  However, the effectiveness of these injections remains a topic of debate. Some studies have shown modest benefits in pain relief and function, particularly in patients with mild to moderate osteoarthritis. However, other research suggests the benefits may be no greater than a placebo effect.  As per the American College of Rheumatology (ACR) and Arthritis Foundation (AF) guidelines from 2019, they conditionally recommend against the use of hyaluronic acid injections for people with osteoarthritis of the knee, and strongly recommend against its use for people with osteoarthritis of the hip or hand. The guidelines suggest that the potential benefits do not outweigh the potential for harm, particularly considering the costs of these injections and the discomfort associated with the procedure.  In general, the primary goals for managing osteoarthritis are to relieve symptoms, improve function, and increase quality of life. Non-pharmacologic treatments such as weight management, physical therapy, and exercise are typically first-line approaches. Pharmacologic treatments like NSAIDs (nonsteroidal anti-inflammatory drugs) are also used. Invasive treatments like injections or surgery are usually considered if other treatments aren't providing adequate relief. | 2 or 3 |
| 22 | Intra-articular (IA) corticosteroids could provide short-term relief for patients with symptomatic osteoarthritis of the knee. | 3 | Are Intra-articular (IA) corticosteroid injection(s) indicated for improving function and quality of osteoarthritis patient? | Intra-articular (IA) corticosteroid injections have been used as a treatment for osteoarthritis to help manage pain and inflammation in the joint. However, their effectiveness in improving function and quality of life is variable and may depend on the specific patient and their circumstances. | 2 or 3 |
| 23 | Platelet-rich plasma (PRP) may reduce pain and improve function in patients with symptomatic osteoarthritis of the knee. | 2 | Are Platelet-rich plasma injection(s) indicated for improving function and quality of osteoarthritis patient? | Platelet-rich plasma (PRP) injections have been proposed as a treatment for osteoarthritis, especially for the knee. PRP is a preparation created from a patient's own blood that is rich in growth factors, which are thought to potentially help in tissue healing and regeneration.  As of my last update in September 2021, the evidence supporting the use of PRP injections for osteoarthritis is still under debate. Some studies have suggested PRP injections may provide pain relief and functional improvement for knee osteoarthritis, while others have found no significant benefit. | 2 or 3 |
| 24 | Denervation therapy may reduce pain and improve function in patients with symptomatic osteoarthritis of the knee. | 2 | Are Denervation therapy indicated for improving function and quality of osteoarthritis patient? | Denervation therapy, also known as radiofrequency ablation (RFA), is a treatment option for chronic pain conditions, including osteoarthritis. It involves the use of heat produced by radio waves to temporarily or permanently interrupt nerve function in a specific area, reducing or eliminating pain signals.  As of my last update in September 2021, denervation therapy has shown some promise in clinical studies for relieving pain from osteoarthritis, especially in the hip and knee. However, these results have been mostly short-term, and there is still much that is not known about the long-term effects and safety of this procedure. | 1 or 2 |
| 25 | Arthroscopy with lavage and/or debridement in patients with a primary diagnosis of knee osteoarthritis is not recommended. | 3 | Are Arthroscopy with lavage and/or debridement indicated for improving function and quality of osteoarthritis patient? | Arthroscopy with lavage (washing out the joint) and/or debridement (removal of damaged tissue) has been used as a treatment for osteoarthritis, especially of the knee. However, the evidence suggests that these procedures may not provide significant long-term benefits for patients with osteoarthritis. | 4 |
| 26 | High tibial osteotomy may be considered to improve pain and function in properly indicated patients with unicompartmental knee osteoarthritis. | 2 | Are High tibial osteotomy indicated for improving function and quality of osteoarthritis patient? | High tibial osteotomy is a surgical procedure that is used primarily in younger, active patients with osteoarthritis primarily affecting one compartment of the knee. The procedure involves cutting and reshaping the tibia (the larger bone of the lower leg) to shift weight from the damaged part of the knee to a healthier area.  As of my last update in September 2021, high tibial osteotomy has shown to provide pain relief and improved function in selected patients with unicompartmental knee osteoarthritis. However, the procedure is generally reserved for younger, more active patients with isolated disease in one compartment of the knee, who have failed non-surgical treatment and are not yet candidates for total knee replacement due to their age or activity level. | 2 or 3 |
| 27 | In the absence of reliable evidence, it is the opinion of the workgroup that the utility/efficacy of dry needling is unclear and requires additional evidence. | 1 | Are dry needling indicated for improving function and quality of osteoarthritis patient? | Dry needling is a technique that involves inserting a needle into a "trigger point" in a muscle in order to relieve pain and improve function. It's often used in the management of musculoskeletal pain, but its effectiveness in osteoarthritis is not well established. | 1 or 2 |
| 28 | In the absence of reliable or new evidence, it is the opinion of the work group not to use free-floating (un-fixed) interpositional devices in patients with symptomatic medial compartment osteoarthritis of the knee. | 1 | Are free-floating (un-fixed) interpositional devices indicated for improving function and quality of osteoarthritis patient? | Free-floating interpositional devices, sometimes called "joint spacer" devices, are used in some cases to manage osteoarthritis, particularly of the knee. These devices are inserted into the joint with the goal of providing cushioning, preserving joint space, and improving function.  As of my last update in September 2021, the evidence regarding the effectiveness of these devices in osteoarthritis management is mixed. Some studies have suggested potential benefits, while others have indicated limited improvement or even complications. Overall, the quality of the evidence is generally considered low, and the use of these devices is usually considered only for specific cases, often after other non-surgical treatment options have failed. | 1 or 2 |
