## supplemental Table 3 for "Assessing the performance of GPT-4 in the filed of osteoarthritis and orthopaedic case consultation"

Supplementery Table 3: 50 orthopaedic cases and response by GPT-4.

|  | Case Description | Further examination | Diagnosis | Treatment |
| --- | --- | --- | --- | --- |
| 1 | The patient is a 39-year-old female who presented with radiating pain in the left upper limb accompanied by numbness in the fingers six months ago. The recovery assessment revealed tenderness in the left neck and shoulder area, positive tests for brachial plexus traction and intervertebral foramen compression on the left side. Additionally, there was reduced skin sensation on the radial aspect of the left upper limb, decreased grip strength, and decreased muscle tone. | Based on the patient's symptoms and physical examination findings, she should undergo cervical spine imaging. Specifically, a magnetic resonance imaging (MRI) scan should be conducted given its superior soft tissue contrast resolution which is particularly helpful in diagnosing conditions such as disc degeneration, herniation, and nerve root compression. This could help reveal any abnormalities in the intervertebral discs, vertebral bodies, spinal cord, or nerve roots. | Considering the patient's clinical presentation and physical evaluation results, her primary diagnosis is likely cervical radiculopathy. This condition is typically caused by nerve root compression in the neck, which may result from factors such as cervical disc herniation, degenerative disc disease, or spinal stenosis. The patient's symptoms of radiating pain, numbness in the left upper limb, and decreased grip strength are indicative of this diagnosis. | The initial treatment for cervical radiculopathy is usually conservative in nature. This includes a combination of rest, physical therapy exercises aimed at strengthening the neck and improving flexibility, non-steroidal anti-inflammatory drugs (NSAIDs) for pain relief, and possibly corticosteroid injections to reduce inflammation around the nerve root. However, if these measures fail to alleviate symptoms or the condition worsens over time, surgical intervention may be necessary. This could involve procedures such as an anterior cervical discectomy and fusion (ACDF), cervical disc arthroplasty, or a posterior cervical laminoforaminotomy, depending on the specifics of the patient's condition. |
| 2 | Patient, female, 45 years old, presented with an open fracture of the lower one-third of the left tibia and fibula. The wound was managed through surgical debridement followed by wound closure and immobilization with a cast. After three months, the patient still reported localized tenderness, and X-ray examination revealed persistent fracture lines with minimal callus formation. Fracture alignment remained satisfactory. | Based on the patient's symptoms and previous X-ray results, she should undergo further imaging to assess the healing status of her fractures. A computed tomography (CT) scan could be ordered for a more detailed assessment of bone detail and alignment, especially as it is often a more accurate tool in evaluating nonunion or delayed union of fractures. Additionally, a bone scan could be considered to evaluate the metabolic activity at the fracture site which can give insight into the healing process. | The primary diagnosis for this patient appears to be delayed union or possibly nonunion of the left tibia and fibula fractures. This is suggested by persistent pain at the fracture site, minimal callus formation and visible fracture lines three months post-injury. | Treatment of delayed union or nonunion depends on various factors including the cause, location, and extent of nonunion. The main goal of treatment is to stimulate bone growth and stabilize the fracture site. Non-surgical interventions such as pulsed ultrasound therapy or bone growth stimulation may be considered initially. However, if these treatments fail or if the nonunion is assessed to be due to infection, mechanical instability or other factors, surgical intervention may be necessary. This might involve bone grafting, internal fixation with plates, screws or intramedullary nails, or external fixation devices. In some cases, if there's an infection present, debridement of the infected bone and tissue would be necessary before bone reconstruction. |
| 3 | A 4-year-old child presented with a history of frequent night crying for the past two weeks, complaining of pain in the left knee joint and reluctance to move the left leg or engage in walking and standing activities. The child had previously visited a local hospital for examination, including X-ray imaging, which did not reveal any significant abnormalities in the left knee. | Considering the age of the child, the symptoms, and the normal X-ray results, it would be advisable to perform a magnetic resonance imaging (MRI) scan of the left hip. The MRI is preferred in this case because it provides excellent contrast resolution and multiplanar imaging capabilities, making it suitable for evaluating soft-tissue abnormalities, bone marrow pathology, or joint disorders. | The primary diagnosis for this patient could possibly be transient synovitis or "irritable hip." This is a common cause of hip pain and limping in children between the ages of 3 and 10. The child's reluctance to bear weight on the affected leg and lack of significant findings on X-ray are typical of this condition. However, other diagnoses, such as Legg-Calvé-Perthes disease (a childhood hip disorder caused by a disruption of blood flow to the ball part of the hip joint) or septic arthritis should also be considered. | In terms of treatment, transient synovitis typically resolves spontaneously with rest and symptomatic treatment, such as pain relief with NSAIDs like ibuprofen. For more serious conditions like Legg-Calvé-Perthes disease or septic arthritis, treatment can range from physical therapy and bracing to surgical intervention, depending on severity of the condition. Regardless, any child with persistent pain and refusal to bear weight requires prompt evaluation and management to prevent long-term complications. |
| 4 | A 60-year-old female patient presented with severe osteoarthritis of the right knee, characterized by flexion contracture of 30° and varus deformity of 25°. The patient underwent total knee arthroplasty and postoperatively received analgesia through a patient-controlled analgesia pump. However, the patient continues to experience pain, numbness, and weakness in the dorsum of the affected limb. | Given the ongoing pain, numbness, and weakness in the limb post-surgery, further imaging studies could be beneficial. An MRI of the right leg may be useful to evaluate for potential nerve damage or compression which could be causing the patient's symptoms. Additionally, X-ray of the right knee could help assess the prosthesis and its alignment. | The primary diagnosis in this case seems to be postoperative neuropathy, possibly due to peroneal nerve injury or another peripheral nerve injury. These types of injuries can occur during total knee arthroplasty due to factors such as direct surgical trauma, nerve stretch, ischemia, or compression from postoperative swelling or hematoma. | Initial management would include medication for pain control, such as NSAIDs or opioids if necessary. Physical therapy could also benefit the patient by improving strength, flexibility, and function of the affected limb. If the patient does not improve or if severe nerve injury is identified on imaging, consultation with a neurologist or neurosurgeon may be warranted for further evaluation and management. This could involve measures ranging from conservative observation, with or without adjuvant therapies like physical therapy and analgesics, to surgical intervention in more severe cases. |
| 5 | A 28-year-old male patient presented with a history of lower back and hip pain for one year, with stiffness upon waking in the morning, and occasional redness and pain in both eyes. Physical examination revealed tenderness at the sacroiliac joint. X-ray examination did not reveal any abnormalities, but the erythrocyte sedimentation rate (ESR) was elevated at 28 mm/h. | Given the patient's symptoms and physical exam findings, further imaging of the sacroiliac joints should be pursued using Magnetic Resonance Imaging (MRI). MRI is more sensitive than X-ray for detecting inflammation or changes in the sacroiliac joints which are often seen in conditions such as ankylosing spondylitis. Besides, it can also show early inflammatory changes before they become evident on X-rays. | The primary diagnosis for this patient appears to be ankylosing spondylitis. This is a type of inflammatory arthritis that predominantly affects the spine and sacroiliac joints. The patient's presenting symptoms of chronic lower back pain, morning stiffness, tenderness at the sacroiliac joint, and occasional redness and pain in the eyes (uveitis), together with elevated erythrocyte sedimentation rate, all support this diagnosis. | The main goals of treatment for ankylosing spondylitis are to reduce symptoms, maintain spinal flexibility, and prevent complications. Non-steroidal anti-inflammatory drugs (NSAIDs) are typically used as first-line treatment to alleviate pain and stiffness. Regular exercise and physical therapy are also critical to help maintain posture and flexibility. In cases where the disease is more advanced or does not respond to NSAIDs, disease-modifying antirheumatic drugs (DMARDs) or biologics might be necessary. Furthermore, the episodes of uveitis should be managed promptly in consultation with an ophthalmologist. |
| 6 | A 6-year-old male patient accidentally noticed a firm protrusion on the inner aspect of the lower end of the right thigh while bathing. The patient did not experience any pain, and knee joint movement was normal. | The recommended imaging test for the child would be an ultrasound, which is non-invasive and doesn't expose the child to radiation. Ultrasound would provide information about the size, shape, location, and characteristics (solid vs liquid) of the protrusion. If further detail is needed - such as evaluation of bone involvement or detailed soft-tissue characterization, an MRI could also be considered. | Based on the provided information, a possible diagnosis might be a benign soft tissue tumor, such as a lipoma or a fibroma. These are common in children and are usually painless. The fact that the protrusion is firm, however, could suggest a more fibrous tumor such as a fibroma or possibly a more serious condition such as a sarcoma. | The treatment plan will depend upon the type of tumor identified. In many cases, if the tumor is benign and isn't causing any symptoms, it can be closely monitored without immediate treatment. However, if the tumor is causing discomfort, or if the diagnosis is uncertain, surgical removal may be necessary. If the tumor is found to be malignant, then a comprehensive treatment plan involving surgery, chemotherapy and/or radiation therapy may be required depending on the specific type and stage of the cancer. |
| 7 | A 21-year-old male patient presented with a one-month history of pain in the lower end of the femur, accompanied by localized swelling on the inner aspect for one week. The affected area showed significant tenderness upon palpation, but the patient did not experience fever or pain during activity. X-ray examination revealed cortical destruction on the medial condyle. | Given the findings from the clinical presentation and X-ray, a further imaging study, such as a magnetic resonance imaging (MRI) scan, should be performed. MRI can provide detailed information about the structure of the femur, including the bone marrow, and surrounding soft tissues. It can help determine the extent of cortical destruction, potential involvement of surrounding tissues and any hidden lesions. | The primary diagnosis for this patient appears to be osteosarcoma. This type of aggressive bone cancer is common in teenagers and young adults and often affects the long bones in the body, such as the femur. The symptoms described by the patient along with the X-ray findings of cortical destruction are indicative of osteosarcoma. | Treatment of osteosarcoma generally involves multimodal therapy combining surgery and chemotherapy. The mainstay of treatment is surgical removal of the tumor, which may involve limb-salvaging surgery or, in more advanced cases, amputation. Adjuvant chemotherapy is typically administered before and after surgery to shrink the tumor and eliminate any remaining cancer cells. The specific treatment plan will depend on the location and stage of the cancer, the patient's overall health, and other factors. |
| 8 | An 8-year-old boy experienced a minor external injury to the left thigh and, on the second day, developed significant localized swelling, severe pain, reluctance to move, and high fever. | Based on the provided case information, this 8-year-old boy should undergo radiographic examinations, specifically an X-ray of the left thigh. This diagnostic test will help to visualize the involved bones and surrounding tissues, potentially revealing any structural damage or changes associated with infection, such as osteomyelitis. Moreover, if necessary, a Magnetic Resonance Imaging (MRI) may be required for a detailed visualization of the soft tissues including muscles and blood vessels. | The main diagnosis that can be inferred from the given symptoms is Acute Hematogenous Osteomyelitis (AHO). AHO is a type of bone infection commonly seen in children, often caused by bacteria (staphylococcus aureus being the most common), which are introduced into the blood stream and reach the bone leading to infection. The injury might have acted as a port of entry for the bacteria. | In terms of treatment, this patient would likely need both medical and surgical intervention. He should be started immediately on empiric antibiotic therapy, typically intravenous, to cover common organisms until culture results are available. Surgery could be needed to drain any abscess and remove any necrotic tissue. After initial management, the patient should be closely monitored and might require physical therapy for rehabilitation. It's also important to communicate with the family about the importance of adhering strictly to the prescribed antibiotics course and follow-up appointments to ensure complete recovery and prevent complications. |
| 9 | A 51-year-old female patient has been experiencing recurrent lower back and leg pain for over six months. She develops intermittent claudication in both legs while walking. The patient complains of numbness and pain in the posterior and lateral aspects of both calves and feet, which is relieved after squatting briefly but does not occur when lying down. There is a slight decrease in sensation in the saddle area and the outer aspect of both legs. Both Achilles reflexes are normal, and dorsalis pedis pulses are present. The patient also has a history of diabetes mellitus for the past 2 to 3 years. | Given the patient's symptoms, we should recommend her to have a Magnetic Resonance Imaging (MRI) scan of the lumbar spine. The MRI can provide detailed images of the spinal cord and surrounding structures, helping to identify any abnormalities such as stenosis, herniation, or other spinal pathologies that might be causing her symptoms. | From the symptoms presented, the primary diagnosis is Lumbar Spinal Stenosis (LSS). This condition is a narrowing of the spinal canal in the lower back, which can compress the nerves and cause symptoms such as pain, numbness, and intermittent claudication. LSS can often occur as a result of wear-and-tear changes in the spine related to osteoarthritis. Additionally, her diabetes might contribute to her symptoms due to diabetic neuropathy, which can cause similar symptoms. | Treatment for Lumbar Spinal Stenosis typically begins with conservative options, including physical therapy, nonsteroidal anti-inflammatory drugs (NSAIDs), and epidural steroid injections. If these measures do not alleviate the symptoms, surgical intervention may be considered, such as decompression surgery or spinal fusion. Her management plan should also include strict control of her diabetes to prevent further nerve damage. It's important to communicate to the patient that while these interventions will help manage the symptoms, they won't cure the condition entirely. Therefore, ongoing management and regular follow-ups are crucial. |
| 10 | A 30-year-old female patient was admitted to the hospital three hours after sustaining a cervical spine injury. Physical examination revealed complete paralysis of all four limbs, with complete loss of sensation, motor function, and reflexes. However, sensation in the acromial region was normal. | Given the history of cervical spine injury and the symptoms described, this patient should undergo immediate imaging studies to assess the extent of the spinal cord damage. I would recommend an emergency Magnetic Resonance Imaging (MRI) scan of the cervical spine. MRI is the imaging modality of choice for spinal cord injuries as it allows visualization of the spinal cord and surrounding tissues, potentially revealing any edema, hemorrhage, or other signs of trauma. In addition, a computed tomography (CT) scan may be indicated if there is a need to evaluate for bony fractures or dislocations. | Based on the physical examination findings, the main diagnosis for this patient suggests a complete Cervical Spinal Cord Injury (SCI). The level of the injury appears to be below the C4 vertebra, since there is normal sensation in the acromial region (innervated by C4), but complete loss of motor function and sensation below that level. | Immediate treatment should focus on stabilizing the patient's neck to prevent further injury. High-dose corticosteroids could be considered within the first few hours after injury to reduce inflammation and swelling around the spinal cord. Surgical intervention may be necessary depending on the nature of the injury - whether there are fractured or dislocated vertebrae that need to be realigned and stabilized. Once the acute phase has passed, the patient will require long-term rehabilitation, including physical and occupational therapy, to manage her symptoms, improve functionality as much as possible, and provide training for activities of daily living given the anticipated permanent paralysis. |
| 11 | A 70-year-old female patient has been experiencing bilateral knee joint pain for over 10 years, with worsening symptoms in the past year, especially after walking long distances. Physical examination reveals medial deviation of both knee joints, and the patellar glide test is negative. X-ray imaging shows narrowing of the medial joint space and the formation of marginal osteophytes. | Given the information provided, and considering the patient's age and radiographic findings, this patient should be recommended to undergo a Magnetic Resonance Imaging (MRI). An MRI would provide detailed images of the soft tissue structures in the knee, including cartilage, tendons, and ligaments that may not be visible on X-ray. This examination can give additional insights into the extent of her condition and help in formulating a precise treatment plan. | Based on the long-term bilateral knee pain, physical and radiographic findings, the primary diagnosis for this patient is Osteoarthritis (OA), likely at a moderate to severe stage. OA is a degenerative joint disease characterized by the breakdown of cartilage and the formation of osteophytes or bone spurs. The medial deviation of the knees, narrowing of the joint space, and presence of marginal osteophytes are all indicative of this condition. | Treatment for osteoarthritis typically includes conservative measures such as pain management with over-the-counter NSAIDs or prescribed medications, physical therapy focused on building strength and improving mobility, weight loss if necessary, and the use of assistive devices like canes or walkers for support. In cases where these measures are ineffective in managing symptoms, surgical procedures like joint replacement surgery may be considered. In addition, it would be beneficial to educate the patient about the importance of lifestyle modifications, such as limiting activities that exacerbate pain, and maintaining a healthy weight to reduce stress on the knee joints. |
| 12 | An 8-year-old male patient with a fracture of the midshaft of the left tibia and fibula for three months. X-ray imaging reveals an oblique fracture of the midshaft of the tibia and fibula, with 60% alignment of the fracture ends, an anterior angulation of 8°, and 1cm overlap. The fracture has already healed. | Given the patient's history and current condition, no further imaging studies may be necessary at this time. As the fracture is already healed, additional imaging would not provide new information or change the management plan. | Based on the provided information, the main diagnosis for this patient is malunion of the fracture. Malunion is a complication of a fracture in which the bone heals in an incorrect position or alignment. This can result in functional impairment or cosmetic deformity. In this case, the oblique fracture of the midshaft of the tibia and fibula has healed with a 60% alignment, an anterior angulation of 8°, and a 1cm overlap. | The treatment approach for malunion depends on the severity of the deformity and the patient's symptoms. If the child does not have significant functional disability, pain, or cosmetic concerns, conservative management with close monitoring might be appropriate. However, if the malunion is causing significant problems, surgical intervention such as osteotomy (cutting and reshaping of the bone) and internal fixation might be necessary to correct the deformity.  It is also crucial to advise the child's parents about the potential risk of future complications due to the improper healing of the fracture. These risks include limb length discrepancy, altered mechanics of the affected limb, and potential early onset arthritis. Regular follow-ups will be needed to monitor the child's progress and manage any arising issues promptly. |
| 13 | A 22-year-old male patient sustained a crushing injury to the right little finger, and it has been three hours since the amputation occurred. The right little finger is completely amputated at the midportion of the first phalanx, while the remaining finger bones are fragmented. There is also partial soft tissue loss. | This patient should be evaluated with a hand x-ray for the remaining portion of the amputated finger and the hand. The purpose of this imaging is to assess the extent of bone damage and fragmentation, which can guide the surgical plan for possible reattachment or reconstruction. | The diagnosis, based on the provided information, is traumatic amputation of the right little finger along with phalangeal fractures. The associated loss of soft tissue further complicates the issue. | In terms of treatment, the first action that needs to be taken is to thoroughly clean and dress the wound to avoid infection. The patient should be administered pain medication and tetanus prophylaxis as necessary. After initial management, a hand surgeon should be consulted immediately to evaluate the need for microvascular surgery for reattachment of the amputated digit. In some cases, reattachment may not be feasible or may lead to less functional outcome, especially when there's severe crush injury or extensive bone fragmentation. If reimplantation is not an option, reconstructive surgery using local flaps or grafts might be considered to cover the wound and maintain the length of the finger if possible. |
| 14 | A 19-year-old female patient presents with left proximal leg pain for the past month without any apparent triggering factors. Physical examination reveals a local bulge with slightly elevated skin temperature and visible varicose veins on the surface. Knee joint mobility is unaffected. X-ray examination indicates "worm-eaten-like destruction" at the proximal end of the tibia. | Given the patient's symptoms and X-ray findings, additional imaging studies should be conducted for further evaluation. Specifically, Magnetic Resonance Imaging (MRI) of the affected area may be advised. An MRI can provide a detailed image of the bone and surrounding tissues, potentially highlighting the extent of bony destruction and its impact on the surrounding structures. | From the provided information, the primary diagnosis is likely a benign bone tumor, possibly an Aneurysmal Bone Cyst (ABC) or Osteoid Osteoma, given her age and the radiographic finding of "worm-eaten-like destruction". ABCs are blood-filled, expansile cystic lesions that can cause pain, swelling, and potential pathological fractures. | For treatment, if the lesion is indeed a benign tumor like an ABC, options could include observation with periodic imaging, sclerotherapy (injecting medicine to shrink the cyst), or surgical curettage (scraping out the lesion) with bone grafting. The choice between these options would depend on factors such as the size and exact location of the lesion, the patient’s symptoms, and the effect of the lesion on the structural integrity of the bone. However, a definitive diagnosis requires biopsy which also helps in ruling out other possibilities including malignancy. It's also important to manage the patient's symptomatic varicose veins with compression stockings and lifestyle modifications. Regular follow-ups are essential to monitor the condition and prevent any complications. |
| 15 | A 60-year-old male patient presents with lower back pain for the past three weeks, without any history of significant trauma. X-ray imaging reveals destruction of the third lumbar vertebra, compression wedge deformity, and normal intervertebral disc space. | Given the patient's age, symptoms and X-ray findings, it would be appropriate to recommend a Magnetic Resonance Imaging (MRI) scan of the lumbar spine. The MRI can provide detailed images of the spinal cord, vertebrae and surrounding structures, potentially revealing any abnormalities such as tumors, infections, or other pathologies that might be responsible for the observed vertebral destruction and deformity. | Based on the provided information, the primary diagnosis for this patient may be a vertebral compression fracture, possibly secondary to osteoporosis or metastatic cancer. The patient's age and the radiographic finding of vertebral destruction and compression wedge deformity without disc space narrowing are suggestive of this condition. | Osteoporotic vertebral fractures are commonly treated with pain management and conservative measures like physical therapy and brace use. In severe cases, minimally invasive surgical procedures like vertebroplasty or kyphoplasty may be considered. However, if the vertebral destruction is due to a pathological process like cancer, then treatment will need to address the underlying condition, potentially involving radiation therapy, chemotherapy, or surgery. It should also be noted that further diagnostic work-up, such as a biopsy or blood tests, may be required to identify the exact cause of the vertebral destruction. Regular follow-ups will be needed to monitor the patient's condition and manage treatment effectively. |
| 16 | A 46-year-old male patient, a long-distance bus driver, has been experiencing recurrent episodes of lower back pain for the past two years. Over the past three months, the pain has started radiating to the left heel. | Considering the patient's symptoms, he should undergo an imaging examination to identify potential abnormalities. Specifically, a Magnetic Resonance Imaging (MRI) scan of the lumbar spine would be recommended. The MRI can provide detailed images of the spinal cord and surrounding structures, helping to detect any spinal pathologies that might be causing his symptoms. | Based on the provided symptoms, primarily the lower back pain radiating to the left heel, the main diagnosis for this patient is likely Sciatica, possibly due to Lumbar Disc Herniation or Degenerative Disc Disease. These conditions can lead to irritation or compression of the sciatic nerve, the longest nerve in the body running from the lower back down to the feet, causing symptoms like lower back pain radiating down to the leg or heel. | Treatment typically involves conservative measures initially, including physical therapy exercises targeting core muscle strengthening, pain relief through non-steroidal anti-inflammatory drugs (NSAIDs), and lifestyle modifications such as weight loss if needed, proper posture, and avoiding sitting for prolonged periods. If these measures are not effective in relieving the symptoms, then epidural steroid injections or surgical intervention may be considered.  In this patient's case, it would also be beneficial to discuss ergonomic modifications such as using a cushion for lower back support while driving or taking regular breaks to stretch. Regular follow-ups should be arranged to monitor the patient's progress and adapt treatment as necessary. |
| 17 | A 32-year-old male patient presented to the hospital with left upper limb dysfunction for the past four months, following a motorcycle accident. Physical examination reveals an inability to perform shoulder abduction and elbow flexion. There is paralysis of the clavicular portion of the pectoralis major muscle, but the latissimus dorsi muscle strength is essentially normal. Movement below the elbow is possible. | Based on the clinical symptoms and history of the trauma, I would recommend an MRI (Magnetic Resonance Imaging) scan of the cervical spine and brachial plexus. This imaging modality can provide detailed images of soft tissues such as nerves and muscles, which may have been damaged in a motorcycle accident. Specifically, this test can help ascertain if there is any nerve root avulsion, nerve compression or other structural abnormalities that could be causing the patient's upper limb dysfunction. | The primary diagnosis for this patient appears to be a brachial plexus injury, specifically affecting the upper trunk which includes the C5 and C6 nerve roots. This damage explains the paralysis of the clavicular portion of the pectoralis major muscle and the inability to perform shoulder abduction and elbow flexion. However, since the patient has retained strength in the latissimus dorsi muscle and can move his arm below the elbow, it is likely that the lower trunk of the brachial plexus (C8-T1) remains intact. | As for the treatment strategy, physiotherapy and rehabilitation exercises should be initiated first to maximize the function of the unaffected muscles and prevent joint stiffness. Pain management, if necessary, could include analgesics or non-steroidal anti-inflammatory drugs. If the MRI reveals severe nerve damage or avulsion, a referral to a neurosurgeon might be needed for surgical exploration and possible nerve repair or grafting. The success of such interventions depends on many factors including the extent and type of nerve injury. Regular follow-ups are essential to monitor the progress of recovery and adjust the treatment plan accordingly. |
| 18 | A 35-year-old male patient has been experiencing pain and swelling on the medial side of the right knee for the past six months. X-ray imaging conducted at an external clinic revealed a translucent area measuring 5cm x 4cm at the proximal medial aspect of the tibia, with a soap bubble-like shadow in the center and enlargement of the bone end. Over the past month, swelling has significantly worsened, with severe nighttime pain and restricted range of motion in the right knee joint. Upon admission, X-ray imaging shows an increase in the size of the lesion at the proximal end of the tibia, disappearance of the soap bubble-like shadow, and the presence of a cloudy shadow infiltrating into the soft tissues. | Given the patient's clinical manifestation, progression of symptoms and radiographic findings, I would recommend a further diagnostic imaging study using MRI (Magnetic Resonance Imaging). This will help to determine the extent of the lesion and any soft tissue involvement. Furthermore, a biopsy might be required for histopathological examination to confirm the diagnosis. | Based on the information provided, the primary diagnosis appears to be Giant Cell Tumor of Bone (GCTB). This is suggested by the age of the patient, site of the lesion, and radiographic findings including 'soap bubble' appearance and an expansive lytic bone lesion. | Treatment plan would be primarily surgical, aiming at complete removal of the tumor to alleviate pain and prevent further enlargement or pathological fracture. The specific type of surgery depends on various factors such as the size and exact location of the tumor, and whether it has spread into nearby structures. Options may include curettage, en bloc resection, or in advanced cases, limb salvage surgery or amputation.  Adjuvant therapy such as denosumab, a monoclonal antibody that inhibits RANKL (a protein essential for the formation, function and survival of osteoclasts which are involved in GCTB), can also be considered. This medication can help reduce tumor size prior to surgery and decrease the risk of recurrence. Postoperative physiotherapy will also be important to regain mobility and strength in the affected knee. Regular follow-ups are necessary to monitor for possible recurrence and manage any complications related to the treatment. |
| 19 | A 29-year-old male patient experienced sudden and severe lower back pain while bending over to lift a bundle of books. The pain was so intense that he was unable to move, and he required assistance from others to return home and rest in bed. The pain persisted the following day. | Given the sudden onset of severe lower back pain associated with a lifting injury, I would recommend an MRI (Magnetic Resonance Imaging) scan of the lumbar spine. The MRI can provide high-resolution images of the spinal structures and help to identify abnormalities such as herniated discs, nerve root compression, or signs of ligament or muscle strain. | Based on the information provided, the primary diagnosis would likely be an acute lumbar disc herniation. This condition occurs when the inner gelatinous material of an intervertebral disc protrudes through the outer layer, potentially pressing on the surrounding nerves and causing severe pain. | The treatment plan for this patient should initially involve conservative management. This includes bed rest in a comfortable position to relieve pressure on the lumbar spine, over-the-counter pain relievers such as nonsteroidal anti-inflammatory drugs (NSAIDs), and physical therapy once the acute pain subsides. |
| 20 | A 67-year-old female patient underwent left knee arthroplasty two weeks ago and has been experiencing pain in the left knee for the past week. Physical examination reveals a body temperature of 36.7℃, a heart rate of 74 beats per minute, and blood pressure of 128/80 mmHg. The left knee is locally swollen and tender, with normal skin color. The skin temperature is slightly higher than the contralateral side. There is localized tenderness on the outer aspect of the inferior pole of the patella, but the patellar glide test is negative. Range of motion of the knee is from 0° to 100°. | Given the patient's postoperative status and recent pain, I would recommend an X-ray of the left knee. This will help to assess the position of the prosthesis and any potential abnormalities that might be causing pain such as loosening of the implant or local bone destruction.  Additionally, I would also suggest obtaining blood tests, particularly inflammatory markers like C-reactive protein (CRP) and erythrocyte sedimentation rate (ESR), to rule out a possible postoperative infection. If suspicion for infection remains high, a joint fluid analysis may be helpful. | Based on the information provided, the primary diagnosis could be postoperative inflammation or early prosthetic joint infection. Both conditions can cause swelling, tenderness, and pain in the surrounding tissues. However, the lack of systemic symptoms such as fever, the slightly elevated skin temperature over the surgical site, and the negative patellar glide test might give more weightage to postoperative inflammation. | The treatment plan should start with conservative management including rest, ice application, elevation of the affected limb, and nonsteroidal anti-inflammatory drugs (NSAIDs) to manage the pain and inflammation.  If infection is suspected based on blood tests or worsening clinical signs, then antibiotics should be started immediately after obtaining appropriate cultures, and consultation with an infectious disease specialist should be considered. In certain cases, surgical debridement or even revision arthroplasty might be necessary.  Regardless of the cause of pain, regular physiotherapy should be continued to improve range of motion and strengthen the muscles around the knee. Regular follow-ups are important to monitor for any signs of complications and to ensure proper healing and recovery. |
| 21 | A 43-year-old male patient has been experiencing lower back pain for the past four years, accompanied by radiating pain in the left lower limb for the past five months. The pain radiates from the lumbar region along the buttocks, posterior aspect of the thigh, and down to the outer edge of the foot. Physical examination reveals a decreased sensation of pain on the posterior and lateral aspects of the right lower leg, as well as the outer edge of the foot. | Based on the patient's clinical history and symptoms, I suggest he undergo a Magnetic Resonance Imaging (MRI) scan of his lumbar spine. This imaging test will help us identify any potential abnormalities or changes in the lumbar vertebrae, intervertebral discs, nerves, and nearby structures that might be contributing to his pain. It can also show whether the nerve roots are compressed or have been damaged due to herniated disc or spinal stenosis. | The patient's symptoms such as chronic lower back pain and radiating pain along the posterior aspect of the thigh and outer edge of the foot seem indicative of Lumbar Radiculopathy, most commonly caused by a herniated disc or spinal stenosis. The decreased sensation of pain in the specified regions further suggests nerve root compression at the L5-S1 level of the lumbar spine. | Our primary treatment goal would be to relieve pain, improve mobility and prevent or minimize future disability. Non-surgical treatments like medication, physical therapy, and epidural steroid injections could be the first step. Medications such as nonsteroidal anti-inflammatory drugs (NSAIDs), muscle relaxants, and narcotics may help reduce pain. Physical therapy exercises aim to strengthen the core muscles supporting the spine which can alleviate pressure on the nerve root causing the pain. Epidural steroid injections can help reduce inflammation around the nerve root. If non-surgical treatments fail to provide relief, surgical intervention like microdiscectomy or laminectomy might be considered, aiming to remove the portion of the disc or bone spur that is pressing on the nerve root. |
| 22 | A 55-year-old male patient was admitted to the hospital two hours after experiencing right hip pain and functional impairment following a car accident. The patient was struck from behind by a low-speed car while crossing the road, resulting in pain in the right hip and limited movement of the affected limb. There are no other discomforts reported and no numbness in the lower limbs. The patient has a history of good health with no previous trauma. Prior to the injury, the affected limb had normal mobility, and there is no history of infectious diseases. Deep tenderness is present in the posterior aspect of the right hip. The affected limb exhibits flexion, adduction, internal rotation, and shortening deformity. Active movement of the hip joint is lost, and passive movement exacerbates the pain with protective muscle spasms. Both knee joints have normal motion, and sensory, motor, and circulatory conditions in the limb are intact. | Based on the patient's history and clinical presentation, it is advisable to perform an X-ray examination of the right hip and pelvis to confirm any possible fractures or dislocations. X-rays are typically the first choice for imaging in cases of trauma as they can give a clear view of the bony structures. If needed, we may also consider a Computed Tomography (CT) scan which can provide detailed images and a better assessment of complex fractures. | From the given symptoms, such as pain, loss of active movement, exacerbated pain during passive movement, and protective muscle spasms, this patient likely suffers from a right hip fracture or hip dislocation. The fact that the limb appears shortened and is held in flexion, adduction, and internal rotation strongly suggests a posterior hip dislocation. | The appropriate treatment would depend on the severity and type of the injury confirmed by imaging studies. For example, if a hip dislocation is confirmed, an immediate closed reduction under sedation or general anesthesia should be performed. This involves manipulating the hip joint back into its normal position. It's crucial to perform this as soon as possible to reduce the risk of complications, like avascular necrosis of the femoral head. If a hip fracture is detected, surgical intervention might be necessary. Depending on the location and extent of the fracture, this could involve internal fixation, partial hip replacement (hemiarthroplasty), or total hip arthroplasty. Post-treatment rehabilitation will play a significant role in regaining mobility and strength. |
| 23 | A 25-year-old male patient presents with lower back pain, fatigue, low-grade fever, night sweats, and weight loss for over a month. The symptoms have worsened over the past three days, leading to hospital admission. The patient previously sought medical attention at an external clinic, where he was diagnosed with "lumbar muscle strain" and prescribed oral pain medication and physical therapy, which provided slight relief. Since the onset of symptoms, the patient has experienced decreased appetite and has lost 3.5kg in weight. Physical examination reveals limited range of motion in the lumbar spine, tenderness at the spinous processes of L4-L5, and normal sensory and motor function in both lower limbs. There is abdominal fullness and mild tension, along with mild tenderness upon gentle palpation on the right lower abdomen. | Given the complex nature of this patient's clinical presentation, I recommend an array of imaging tests to identify the root cause. A Magnetic Resonance Imaging (MRI) scan of the lumbar spine can provide detailed information about any abnormalities in discs, vertebrae, or soft tissues that may be contributing to his lower back pain. Given that he also presents with symptoms such as fatigue, fever, night sweats, and weight loss, these systemic symptoms indicate a possible underlying infectious or neoplastic condition. To explore this further, Abdominal Ultrasound or CT scan could help detect any abdominal pathologies potentially linked to his right lower abdominal tenderness and fullness. | The clinical picture provided indicates potential Lumbar Spondylodiscitis, an infection of the intervertebral disc space and adjacent vertebrae, which is consistent with his back pain, tenderness at L4-L5, and systemic symptoms. His right lower abdominal tenderness may be indicative of an intra-abdominal process such as appendicitis or possibly an abscess formation if there's an ongoing systemic infection. | Treatment will depend on definitive diagnosis. If lumbar spondylodiscitis, antibiotic therapy, possibly combined with surgical intervention might be required. If an intra-abdominal process is confirmed, it might need surgical attention. Regardless, this patient needs to be kept under close observation due to the severity and complexity of his symptoms. |
| 24 | A 37-year-old male patient was admitted to the hospital 24 hours after a motor vehicle accident, where he lost control of the car and collided head-on with an obstacle. The patient experienced impaired motor and sensory function in all four limbs following the injury. Vital signs have remained stable since the injury, and the patient is conscious and alert. X-ray imaging of the cervical spine reveals loss of physiological curvature and incomplete segmental fractures of the C5 and C6 vertebral bodies, with no evidence of fractures or dislocations in other spinal segments. | Given the patient's clinical manifestations and X-ray findings, I would recommend further imaging studies including a computed tomography (CT) scan and magnetic resonance imaging (MRI) of the cervical spine. The CT scan can provide more detailed images of fractures and is especially helpful in assessing the degree of bony injury. An MRI scan can visualize any damage to soft tissues, such as ligaments, discs, and especially the spinal cord, which may have occurred due to the trauma. | The patient's symptoms are indicative of a traumatic spinal cord injury at the cervical level given the impaired motor and sensory function in all four limbs (quadriplegia/ tetraplegia). The X-ray findings confirm evidence of trauma with loss of physiological curvature and incomplete segmental fractures of C5 and C6 vertebral bodies. | Treatment for this patient would be multidisciplinary and involve several stages. Initially, measures should be taken to stabilize his spinal cord injury to prevent further damage. This might include immobilization, corticosteroid medication to limit inflammation, or surgery to decompress the spinal cord and/or stabilize the fractured vertebrae. Following the acute stage, rehabilitation will be crucial for helping the patient recover the maximum possible function and adjust to life after injury. This may involve physical therapy, occupational therapy, vocational therapy, counseling, and other supportive services. It is important to note that recovery from a spinal cord injury can be prolonged and each person's recovery potential can be different. |
| 25 | A 30-year-old male patient who fell from a height presents with abdominal tenderness and rebound tenderness. The patient also reports significant pain and deformity in the right thigh. Despite these symptoms, the patient is conscious and alert. | Given the patient's symptoms and the mechanism of injury, I would recommend a series of imaging studies. An abdominal computed tomography (CT) scan would be appropriate to assess the extent of the intra-abdominal injuries and to identify any potential internal bleeding. As for the right thigh pain and deformity, an X-ray of the femur should be performed initially to evaluate possible fractures. | The abdominal tenderness and rebound tenderness could suggest intra-abdominal trauma, possibly involving organ injury or internal bleeding. The reported significant pain and deformity in the right thigh may indicate a fractured femur. | Treatment will depend on the findings from the imaging studies. If internal abdominal injuries are confirmed, management might range from conservative monitoring to surgical intervention, depending on the severity of the injuries. A suspected femur fracture would typically require orthopedic consultation for possible surgical repair with intramedullary nailing or plating, depending on the type and location of the fracture. Post-surgery, the patient would benefit from physical therapy as part of his rehabilitation to regain strength and mobility in the injured limb. |
| 26 | A young athlete presents with patellar pain and weakness in the right knee for the past three months. The patient experiences pain during knee flexion and extension and has difficulty going up and down stairs. Physical examination reveals tenderness along the edge of the patella, a positive patellar grinding test, and mild atrophy of the quadriceps muscle. Plain X-ray imaging of the right knee does not show any abnormalities, but nuclear scanning reveals localized radiotracer uptake in the patella. | Based on the patient's symptoms and physical examination findings, the next imaging study that should be performed is magnetic resonance imaging (MRI) of the right knee. Although the plain X-ray did not show any abnormalities, MRI can provide more detailed information about the soft tissues surrounding the patella, including the tendons, ligaments, cartilage, and muscles. It can help identify any potential structural abnormalities or inflammation that may be causing the patellar pain and weakness. | The main diagnosis for this patient based on the clinical presentation and nuclear scanning results is patellar tendinopathy, also known as jumper's knee. This condition is characterized by pain and dysfunction in the patellar tendon due to repetitive overuse or strain. The localized radiotracer uptake in the patella seen on nuclear scanning indicates increased metabolic activity in the area, which is consistent with inflammation or degeneration of the patellar tendon. | Treatment options for patellar tendinopathy typically include a combination of conservative measures and rehabilitation exercises. These may include rest, ice, compression, elevation (RICE), non-steroidal anti-inflammatory drugs (NSAIDs) for pain relief, physical therapy to strengthen the quadriceps and improve biomechanics, and modification of activities or sports techniques to reduce stress on the patellar tendon. In some cases, more advanced treatment options such as extracorporeal shockwave therapy or platelet-rich plasma injections may be considered. Surgical intervention is rarely necessary but may be considered in severe or refractory cases. Regular follow-up and monitoring of symptoms and response to treatment are important to ensure proper management of the condition. |
| 27 | A 29-year-old male patient with a transverse fracture of the midshaft of the right humerus underwent open reduction and internal fixation with a plate four months ago. Follow-up X-ray examination shows good alignment and apposition of the fracture, with no evidence of loosening in the internal fixation. The fracture line is clearly visible without significant callus formation. | From the presented information, although the alignment, apposition of the fracture, and internal fixation appear satisfactory on X-ray, the lack of significant callus formation four months post-surgery suggests that this patient may be experiencing delayed union.  Delayed union is a temporary condition in which adequate healing has not occurred in the expected time frame. This can occur for several reasons, including poor blood supply, infection, inadequate immobilization, or the patient's own health status (e.g., smoking, diabetes).  To further evaluate this situation, an MRI could be helpful to assess soft tissue around the fracture site and check for any signs of infection or lack of blood flow. It can also provide additional detailed images of bone marrow, which can help evaluate the process of bone healing. | The primary diagnosis for this patient would be delayed union of the right humerus fracture. The treatment could include ensuring optimal conditions for bone healing. This can involve revising the surgical approach if necessary, controlling any possible infections, managing underlying medical conditions, and adjusting lifestyle factors (such as encouraging smoking cessation, if applicable). | In addition, physical therapy may be beneficial to encourage mobilization and prevent joint stiffness, while being cautious not to apply excessive load to the affected limb. It's important to closely monitor the patient's progress through regular follow-up appointments and additional imaging studies as needed. If there is no improvement after these measures, further surgical management such as bone grafting might be considered. |
| 28 | A 37-year-old male patient presents with pain and swelling in the lower end of the left femur, along with mild restriction of knee joint movement for the past seven days. Physical examination reveals a localized, tender prominence on the outer side of the distal femur, with normal skin temperature. X-ray examination shows an eccentrically growing bone resorption lesion on the lateral condyle of the femur, with outward cortical expansion and no periosteal reaction. | Given the patient's presentation and radiographic findings, further imaging studies are warranted to better define the lesion. A magnetic resonance imaging (MRI) study of the left knee should be performed next. MRI is typically the best imaging modality for characterizing bone lesions. It can provide detailed information about the lesion itself, including its size, composition, and whether or not it involves surrounding soft tissue structures. Furthermore, MRI can help identify any potential pathological fractures or associated marrow edema. | The primary diagnosis for this patient, based on clinical evaluation and X-ray findings, seems to suggest a benign bone tumor, possibly a chondroblastoma or enchondroma. These tumors may cause localized pain, swelling and can lead to eccentrically located bone resorption with outward cortical expansion. | Treatment options depend on the exact type and stage of the tumor. For benign conditions like chondroblastoma or enchondroma, curettage (surgical scraping of the lesion) followed by bone grafting is often the treatment of choice. If the tumor is more aggressive or malignant, surgical resection might be required. In either case, referral to an orthopedic oncologist would be advisable for further evaluation and management.  The final treatment plan will be determined after a thorough discussion with the patient considering the benefits and risks of each option. Regular follow-up visits will be necessary to monitor the healing process, manage potential complications and detect any recurrence early. |
| 29 | A 44-year-old female patient complains of pain and snapping at the base of the palm on the volar aspect of the right thumb for the past year, with worsening symptoms over the past two weeks. Physical examination reveals a palpable nodule at that site, which is tender to touch. Snapping can be felt when flexing and extending the thumb. | Given the patient's symptoms and physical examination findings, an ultrasound of the right thumb should be undertaken. Ultrasound can provide real-time imaging of the musculoskeletal structures in the thumb, including tendons, pulleys, and soft tissue nodules. This could enable us to visualize any abnormalities such as a stenosing tenosynovitis or a tendon sheath ganglion that may be causing the triggering or snapping of the thumb. | The primary diagnosis for this patient, based on her clinical presentation, is trigger thumb, also known as stenosing tenosynovitis of the thumb. This condition occurs when inflammation or nodular thickening of the flexor tendon or its sheath at the A1 pulley level interferes with the normal gliding mechanism during thumb motion, leading to triggering or snapping and potential locking in a bent position. | Treatment usually starts with conservative measures such as rest, splinting, non-steroidal anti-inflammatory drugs (NSAIDs) for pain relief, and potentially a corticosteroid injection into the tendon sheath to reduce inflammation and swelling. If these measures are not effective, surgical intervention might be considered, which typically involves releasing the constriction by dividing the affected part of the tendon sheath (A1 pulley). Physical therapy may also be recommended post-surgery to regain full function of the thumb. |
| 30 | A 3-year-old male patient exhibits a gait pattern with toe-walking and landing on the lateral aspect of the foot, resulting in an unstable gait. Physical examination reveals foot dorsiflexion, forefoot adduction, inversion, and internal rotation deformities, but normal skin sensation. | Considering the child's age and the symptoms described, it would be advisable to carry out a series of X-rays on both lower limbs and particularly focusing on the foot and ankle. These X-rays should be taken in different positions, such as weight-bearing and non-weight-bearing, to provide a comprehensive view of any potential deformities or abnormalities. | The primary diagnosis for this patient, based on his clinical presentation, is Clubfoot, also known as Congenital Talipes Equinovarus (CTEV). This condition is characterized by four main components: midfoot cavus, hindfoot varus, forefoot adduction and equinus at the ankle. It typically presents at birth and complicates walking if not treated early. | Treatment for clubfoot is typically initiated as soon as possible and often involves a non-surgical method known as the Ponseti method. This includes gentle manipulation and casting of the foot, which is changed weekly over several weeks, gradually correcting the position of the foot. After this period, the child usually undergoes a minor procedure to release the Achilles tendon (tenotomy), followed by bracing to maintain the correction. |
| 31 | A 40-year-old female presented with bilateral hip and leg pain during continuous walking, necessitating intermittent squatting for the past two years. Initially, she was able to walk continuously for half an hour, but over time, the duration of her symptom-free intervals gradually decreased. Currently, she experiences symptoms after walking just 200 meters. Interestingly, she remains asymptomatic when lying flat. On physical examination, tenderness was observed at the L4-L5 intervertebral space without any radiation. Straight leg raising test showed a full range of motion bilaterally at 70°. Sensation and muscle strength in both lower extremities were normal. | Given the patient's symptoms and physical examination findings, further imaging studies are needed to provide a more definitive diagnosis. A lumbar spine MRI should be performed. MRI allows for the visualization of soft tissues including intervertebral discs, spinal cord, nerve roots, and muscles, it can help detect any potential disc herniation, spinal stenosis or other abnormalities that may be causing the patient's bilateral hip and leg pain. | The primary diagnosis for this patient based on her clinical presentation is neurogenic claudication possibly due to lumbar spinal stenosis. Lumbar spinal stenosis is a condition where narrowing of the spinal canal occurs, which can place pressure on the spinal cord and nerves within, leading to symptoms such as pain, numbness, or weakness in the legs. | As for the treatment plan, initial management of lumbar spinal stenosis typically involves conservative measures such as physical therapy for improving flexibility, strength and balance, non-steroidal anti-inflammatory drugs (NSAIDs) for pain relief, epidural steroid injections for reducing inflammation and alleviating pain, and lifestyle modifications like weight loss and quitting smoking to reduce pressure on the lower back. If these measures fail to alleviate symptoms, surgical options like decompressive laminectomy or lumbar fusion surgery might be considered. |
| 32 | A 60-year-old male presented with a neck mass for two years and hoarseness of voice for the past four months. Two months ago, he underwent a total thyroidectomy due to a thyroid tumor, as confirmed by the pathology report. Following the surgery, he accidentally slipped and fell on his buttocks while getting out of bed, resulting in low back pain. Over the past two weeks, the back pain has worsened to the point where he is unable to mobilize from bed. A lateral X-ray of the lumbar spine revealed compression and flattening of the L3 vertebral body with uneven density. The right pedicle of the vertebra appears to be absent, while the intervertebral space appears normal. | Given the patient’s history of a thyroid tumor, his recent fall, and radiographic findings, the next imaging study to be performed should be magnetic resonance imaging (MRI) of the spine. This would help to further investigate the vertebral changes seen on the X-ray and to evaluate for possible metastasis from the thyroid cancer. MRI provides high-resolution images and is more sensitive in detecting bone metastases compared to an X-ray. | The primary diagnosis for this patient, based on his clinical presentation and X-ray findings, is likely metastatic thyroid cancer to the lumbar vertebrae. The symptoms of back pain and immobility, together with the abnormal findings on the X-ray including compression, flattening, absent pedicle, and uneven density of the L3 vertebra, can signify destructive metastatic lesions. | Given this diagnosis, treatment options would often involve a multidisciplinary team approach including endocrinologists, oncologists, radiation therapists, and orthopedic surgeons. Treatment plans are individualized and may include systemic therapy such as radioactive iodine, targeted therapies or chemotherapy, local therapy such as external beam radiation to relieve pain and prevent pathological fractures, and surgical intervention if indicated for spinal stabilization. |
| 33 | A 66-year-old female patient has been experiencing recurrent headaches and dizziness over the past six months. This morning, she suddenly felt vertigo, tinnitus, and nausea with vomiting when she turned her head quickly. She subsequently fell to the ground but experienced relief after approximately two minutes. The patient reported having had similar episodes twice in the past. X-ray images revealed posterior osteophyte formation and significant narrowing of the intervertebral space at the C5-C6 vertebral level. | Given the patient's symptoms and X-ray findings, further imaging studies are necessary to gain a more comprehensive understanding of her condition. A cervical spine MRI should be performed next. This will allow for detailed examination of the soft tissues including the spinal cord, nerve roots, and intervertebral discs. It can help identify potential causes of vertigo and other neurological symptoms such as disc herniation, spinal stenosis, or foraminal narrowing due to osteophyte formation. | The primary diagnosis for this patient, based on her clinical presentation and X-ray findings, is likely cervical spondylosis with possible vertebrobasilar insufficiency. Cervical spondylosis refers to degenerative changes in the cervical spine causing narrowing of the intervertebral space and formation of osteophytes. These changes may potentially impinge upon the vertebral arteries, leading to insufficient blood flow to the posterior brain (vertebrobasilar insufficiency), which can cause symptoms such as dizziness, vertigo, tinnitus, nausea, and vomiting. | Treatment usually begins with conservative measures, including physical therapy for improving posture, flexibility, and strength, pain management with non-steroidal anti-inflammatory drugs (NSAIDs), and lifestyle modifications. If these measures are unsuccessful, surgical intervention might be considered depending on the severity of the spinal cord or nerve root compression and the patient's overall health status. |
| 34 | A 3-year-old male child presented with sudden crying and refusal to use his right upper limb when his mother tried to pull him up the stairs. | Based on the history provided, it would be appropriate to perform an X-ray imaging of the child's right upper limb. This can help us identify any potential fractures, dislocations, or other injuries that could have been caused by the pulling action. | The primary diagnosis for this situation, based on the described clinical presentation, is likely Nursemaid's Elbow, also known as Radial Head Subluxation. This condition is commonly seen in young children and occurs when the radial head (part of the arm bone) slips out of its normal position at the elbow joint. It typically happens following a sudden pull on the extended pronated arm, just as described in this case. | Treatment usually involves a simple reduction procedure performed by a healthcare professional. This is typically done without the need for anesthesia and results in immediate relief of symptoms in most cases. The child should be able to use his arm normally shortly after the reduction. After the procedure, instructions will be given to avoid pulling or lifting the child by the arm to prevent recurrence of the subluxation. Regular follow-up visits may be necessary to ensure proper healing and function of the elbow joint. |
| 35 | A 51-year-old male patient presented with pelvic pain following a car accident. He is unable to urinate spontaneously and experiences bladder distension. There is a small amount of bloody discharge from the urethral orifice. His blood pressure is measured at 80/60 mmHg. A digital rectal examination revealed blood-stained gloves. | Based on the clinical presentation, urgent imaging is required to assess the extent of damage. A pelvic X-ray should be performed initially to identify any potential fractures or dislocations. This should be followed by a computed tomography (CT) scan of the abdomen and pelvis with intravenous contrast to further evaluate for any intra-abdominal or pelvic injuries. CT scan has the advantage of being able to visualize soft tissues, bones, blood vessels, and other structures within the body and can help detect any evidence of internal bleeding, organ rupture, or bladder injury. | The patient's symptoms and examination findings suggest the possibility of a pelvic fracture with accompanying urethral injury. The low blood pressure could be indicative of shock due to internal bleeding associated with the trauma. Blood at the urethral meatus and inability to urinate point towards possible urethral disruption. | Immediate treatment will involve stabilizing the patient's condition starting with addressing the hypotension, which might require intravenous fluids or blood transfusion as necessary. Pain management will also be crucial with appropriate analgesics. Urethral catheterization should be avoided until a urethral injury has been ruled out since it can exacerbate the damage.  Once stabilized, orthopedic surgery may be required to fix any bone fractures, and urologic consultation is needed for management of the suspected urethral injury. Depending upon the severity of the urethral injury, conservative management with a suprapubic catheter or surgical intervention may be considered.  Regular follow-up appointments will be necessary to monitor the patient's response to treatment and to manage any potential complications. |
| 36 | A 22-year-old male patient presented with swelling, pain, and functional impairment in his left lower leg following a sports-related injury. The patient reported experiencing sudden and intense pain in his left lower leg while rapidly changing direction to chase a soccer ball one hour prior to presentation. As a result, he was unable to move and required urgent transportation to the hospital. | Based on the patient's history and symptoms, the recommended imaging study in this case would be an ultrasound of the lower extremity to assess for any soft tissue injuries. Additionally, MRI of the left lower leg should be considered as it can provide a more detailed evaluation of the muscle, tendon, ligament, and bone involvement. | The primary diagnosis for this patient, based on his clinical presentation, is likely an acute Achilles tendon rupture. This is a common injury seen in athletes, particularly those involved in sports that require sudden changes in direction or speed, such as soccer. | Treatment typically depends on several factors including the patient’s age, activity level, and the severity of the injury. Non-operative management involves immobilization in a cast or boot with the foot in a position of plantar flexion, followed by physical therapy for rehabilitation. Surgical repair is often recommended for young, active individuals due to a lower risk of re-rupture compared with non-operative treatment. |
| 37 | A 26-year-old female patient presented with swelling, pain, bleeding, and limited mobility in her right lower leg following a car accident. On examination, her vital signs were stable. A 13 cm laceration was observed on the upper-middle section of the skin of her right lower leg, with an exposed fracture end of the tibia. Although there was significant soft tissue contusion, no obvious tissue loss was noted, and bleeding was minimal. The dorsalis pedis artery was palpable. | Firstly, in terms of imaging studies, it's crucial to perform an X-ray examination of the right lower leg. This radiographic examination will help us assess the type and extent of the tibial fracture and any possible involvement of the fibula. Also, it is essential to evaluate the joint above (knee) and below (ankle) the fracture site for any associated injuries.  Secondly, a CT scan might be recommended depending on the complexity of the fracture observed in the X-ray. The CT scan can provide more detailed information about the fracture pattern, comminution, and intra-articular extension, which could influence the surgical plan. | According to the clinical presentation and physical examination, the primary diagnosis for this patient is an open fracture of the tibia, more specifically, it likely falls under Gustilo and Anderson type II or IIIA open fracture given there is no significant soft tissue loss but involves a larger wound with fracture contamination. | The treatment should follow the guidelines for open fractures. First, antibiotic prophylaxis should be initiated immediately, preferably within three hours of injury, to prevent infection. As the patient's tetanus immunization status is not known, tetanus prophylaxis should also be considered.  In addition, the patient requires urgent surgical intervention. The initial management should involve thorough irrigation and debridement of the wound in the operating room, followed by fracture stabilization. Depending on the specific characteristics of the fracture, this may be accomplished through external fixation, intramedullary nailing, or plate fixation.  Finally, further wound care including possible secondary wound closures or skin grafting may be required, as well as long-term follow-up for rehabilitation and monitoring for potential complications such as infection, nonunion, malunion, or post-traumatic arthritis. |
| 38 | A 12-year-old female patient, a student, presented with a one-week history of fever accompanied by right lower leg pain for the past three days. The patient had experienced fever and sore throat after being exposed to cold temperatures one week ago. The parents self-administered over-the-counter cold medications (specific name unknown) without seeking medical attention. Over the past three days, the patient developed chills, high fever reaching up to 40℃, and severe pain in the right lower leg, which limited her movement. On examination, the temperature was 38.8℃, respiratory rate was 30 breaths per minute, heart rate was 110 beats per minute, and blood pressure was 100/50 mmHg. Specialized examination revealed no spinal curvature or tenderness. The right knee joint was partially flexed and had limited range of motion. The floating patella test was negative. There was significant tenderness in the upper part of the right lower leg, but no apparent swelling. The local skin temperature was elevated with slight redness, without visible varicose veins. No abnormalities were found in the other limbs. Blood tests showed a white blood cell count of 19×10^9/L, with 93% neutrophils; hemoglobin level of 92 g/L; platelet count of 213×10^9/L; and an erythrocyte sedimentation rate of 30 mm/h. | Based on the patient's presenting symptoms, I would recommend an X-ray of the right lower leg and knee joint as an initial imaging study to rule out any osseous abnormalities or signs of osteomyelitis. Additionally, a Magnetic Resonance Imaging (MRI) might be necessary for a more detailed assessment of soft tissue involvement, which is often missed on X-rays. | The clinical findings of high fever, localized pain along with the raised count of white blood cells and high proportion of neutrophils suggest an infection. The right lower leg pain could be suggestive of osteomyelitis, which is a bone infection usually caused by bacteria or other germs. This can occur after a minor trauma, spread from an infection in a nearby tissue, or reach the bone via the bloodstream. | As for treatment, if osteomyelitis is indeed confirmed, it typically involves several weeks of intravenous antibiotic therapy, followed by a few weeks of oral antibiotics. It's also important to manage symptoms and improve function. For pain management, over-the-counter pain relievers such as acetaminophen or ibuprofen may be used. In severe cases or when the infection does not respond to antibiotics, surgery may be required to remove infected tissue and repair any damage to the bone. |
| 39 | Immediate intervention is warranted in a 20-year-old male patient who had previously suffered from acute purulent osteomyelitis of the right femur six months ago. The patient exhibited improvement after treatment; however, the presence of a sinus tract with occasional discharge of scanty yellowish purulent fluid has persisted in the affected area. Recently, for the past four days, the sinus tract has closed, but the patient has developed high fever and significant local tenderness accompanied by erythema and swelling. X-ray imaging demonstrates the presence of residual necrotic bone with well-formed sequestrum formation. Given these findings, prompt action is necessary. | In terms of imaging, the patient has already undergone an X-ray which revealed the presence of necrotic bone. However, it would be appropriate to proceed with an MRI to assess the extent of the soft tissue involvement and to detect any possible abscess formation. Additional imaging modalities such as a bone scan or, in some cases, a CT scan may also be recommended depending on the individual case scenario. | For this patient, given his history of acute purulent osteomyelitis and the current presentation of recurring symptoms and X-ray findings, we need to consider chronic osteomyelitis. The closure of the sinus tract, along with recurrent signs of infection including fever, tenderness, swelling, and erythema, coupled with radiographic evidence of sequestrum (dead bone that has separated from living bone), are indicative of such a diagnosis. | As for treatment, this will largely depend on the severity of the condition, but generally involves both surgical and medical management. Surgical intervention is often necessary to debride necrotic bone and eliminate any abscesses. This can involve surgical drainage, debridement of the sequestrum, and possibly local antibiotic delivery with bone grafting if there's significant bone loss.  After surgery, the patient will typically require prolonged antibiotic therapy, guided by culture and sensitivity results. It's crucial to understand that treating chronic osteomyelitis can be a long process, often requiring weeks to months of antibiotics, and sometimes multiple surgical procedures. Appropriate physiotherapy should also be included in the plan to maintain joint mobility and muscle strength.  Prompt and aggressive treatment of this patient is critical to avoid serious complications like pathological fractures, septicemia, or even amputation, especially considering the patient's young age. |
| 40 | A 40-year-old male presents with pain at the distal end of the femur and mild restriction of knee joint mobility. Physical examination reveals a localized bulge, tenderness, and slightly elevated skin temperature on the lateral aspect of the distal femur. X-ray imaging shows eccentrically growing bone resorption lesions on the lateral condyle of the femur, accompanied by outward bulging and thinning of the cortex, without periosteal reaction. | Given the patient's clinical presentation and radiographic findings, the recommended imaging examination would be a Magnetic Resonance Imaging (MRI) scan. MRI would allow for better characterization of the bone lesion and assessment of any soft tissue involvement. It is particularly useful in evaluating the extent of the lesion, detecting any underlying pathological fracture, and ruling out potential invasion into joint space or adjacent neurovascular structures. | From the information provided, the main diagnosis to consider is a benign bone tumor, such as an osteochondroma. These typically present with asymptomatic bony growths that can cause pain and hinder joint mobility when they press on nearby nerves or tissues. The eccentric growth pattern, outward bulging, and cortical thinning without periosteal reaction observed on X-ray are suggestive of this condition. | The treatment approach depends on several factors, including the size and location of the tumor, the patient's symptoms, and whether the tumor is growing. If the osteochondroma is small, not causing significant symptoms, and not growing, observation might be all that's needed. However, if the patient presents severe pain, limited mobility, or if the tumor is growing or threatening nearby structures, surgical excision could be considered. Preoperative planning should involve a multidisciplinary team, including orthopedic oncology specialists, to ensure optimal outcomes. Remember to provide adequate post-operative care and regular follow-up to monitor for recurrence or malignant transformation, although these are relatively rare in osteochondromas. |
| 41 | A 40-year-old male with a history of long-term corticosteroid use due to a skin disease presents with right hip joint pain, limited mobility, and approximately 2cm shortening of the right lower limb over the past six months. | Given this patient's history and presentation, the recommended imaging examination would be an X-ray of the right hip. This will help determine any changes in joint space, structural deformities, or presence of bone lesions. Based on the symptoms and their progression, it may also be beneficial to perform a Magnetic Resonance Imaging (MRI) scan. The MRI can provide more detailed information about soft tissues, cartilage, and the extent of potential bone damage not visible on an X-ray. | The main diagnosis to consider for this patient is avascular necrosis (AVN) of the femoral head, also known as osteonecrosis. AVN occurs when blood flow to a section of bone is interrupted, causing the bone cells to die which can lead to pain and decreased mobility within the joint. Long-term use of corticosteroids is a well-known risk factor for AVN. | The treatment of AVN varies according to the severity and stage of the disease at diagnosis. Initially, conservative measures such as physical therapy, weight-bearing restrictions, analgesics, and bisphosphonates may be used to manage pain and slow down the progression of the disease. However, if these measures are ineffective or the disease is advanced, surgical intervention may be considered. Surgery could involve core decompression to reduce intraosseous pressure and promote revascularization, or joint replacement in severe cases with advanced collapse of the femoral head. It is also crucial to address the underlying cause, in this case, the long-term use of corticosteroids, by exploring alternative treatment options for the patient’s skin condition under the care of a dermatologist. |
| 42 | A 15-year-old female patient with a history of right distal humerus fracture five years ago presents with suboptimal reduction and subsequent cubitus valgus deformity. Six months ago, she started experiencing numbness along the ulnar aspect of her right hand. | The recommended imaging examinations for this patient would be an X-ray of the right elbow, and a Nerve Conduction Study (NCS) combined with Electromyography (EMG). The X-ray will help in assessing the degree of deformity, determining the status of the previously fractured site, and planning any future surgical intervention if necessary. Due to her symptoms of numbness, the NCS and EMG would be useful in confirming any nerve damage or entrapment, specifically that of the ulnar nerve. | Considering the history and presentation, the main diagnosis is likely cubitus valgus deformity resulting from improper healing of the distal humerus fracture, subsequently leading to ulnar nerve compression, often referred to as "cubital tunnel syndrome". | The treatment for this condition is multifaceted. Initially, physiotherapy and non-steroidal anti-inflammatory drugs (NSAIDs) can be used to manage the symptoms. Bracing or night splints can also be implemented to alleviate symptoms by preventing flexion of the elbow during sleep, which can exacerbate nerve irritation. However, given the long-term presence and progression of symptoms, surgical intervention may be warranted. This could involve corrective osteotomy to realign the bones and relieve pressure on the ulnar nerve. In severe cases of nerve entrapment, ulnar nerve transposition might be needed. Postoperative physiotherapy would be crucial for functional recovery. Regular follow-up is also essential to monitor signs of improvement or complications. |
| 43 | A 58-year-old female fell while walking outside and experienced severe pain in her right hip. She was unable to bear weight on her right lower limb and required assistance to walk. Physical examination reveals an externally rotated and abducted deformity of the right lower limb, with shortening. Tenderness is present along the longitudinal axis of the right lower limb, and there is a positive test for greater trochanteric prominence. | Based on the patient's history of a fall, her clinical manifestation and physical examination findings, the recommended imaging examination would be an X-ray of the right hip in both anteroposterior (AP) and lateral views. This will allow for a comprehensive evaluation of the hip joint to identify any fractures, dislocations or other abnormalities. | The main diagnosis to consider is a right femoral neck fracture which often occurs following a low-energy trauma, such as a fall, especially in older individuals with risk factors such as osteoporosis. The clinical presentation of severe pain, inability to bear weight, abnormal limb position, and positive test for greater trochanteric prominence are highly suggestive of this condition. | Treatment for femoral neck fractures depends on several factors including the patient’s age, overall health status, and the type and location of the fracture. Non-displaced fractures might be treated with internal fixation using screws. However, displaced fractures in older patients usually require arthroplasty, either a hemiarthroplasty or a total hip replacement, due to high rates of non-union and avascular necrosis associated with these types of fractures.  Regardless of the specific surgical procedure performed, postoperative care should include pain management, prevention of deep vein thrombosis, and early mobilization with physical therapy to restore function and independence. Additionally, addressing underlying risk factors such as osteoporosis is essential for prevention of future fractures. |
| 44 | Following a sudden brake of a public bus, the patient's head and face struck the backrest of the front seat, resulting in restricted neck movement and flaccid paralysis of all four limbs. Sensory and motor functions are lost below the level of the sternal body plane, and the patient is unable to urinate voluntarily. | Given the patient's symptoms and history, the recommended imaging examinations would be a computed tomography (CT) scan and magnetic resonance imaging (MRI) of the cervical spine. This will help in identifying any bone fractures or dislocations and assessing for soft tissue damage including spinal cord injury. | From the provided information, the main diagnosis to consider is a traumatic spinal cord injury, specifically at the cervical level. The sudden impact on the head and neck could have resulted in abrupt displacement or fracture of the cervical vertebrae, leading to acute spinal cord compression or transection. This is supported by the immediate onset of quadriparesis (weakness in all four limbs), loss of sensory and motor functions below the chest, and inability to urinate voluntarily, which indicates disruption of autonomic functions. | The treatment of such severe spinal cord injuries is a complex process involving multiple specialties. Initially, it's crucial to stabilize the patient's vital signs and prevent secondary injuries. High-dose corticosteroids may be administered within the first few hours following the injury to reduce inflammation and secondary neurological damage.  Once the patient is stabilized, surgery is often required to decompress the spinal cord and fixate the unstable spinal segments. This could involve removal of bone fragments, disc material, or fractured vertebral bodies that are compressing the spinal cord (decompressive laminectomy or corpectomy), followed by spinal fusion and internal fixation. |
| 45 | A 19-year-old student has been experiencing pain in the distal end of the right femur for the past three months, with worsened symptoms at night. Examination reveals a localized bulge on the medial aspect of the distal end of the right femur, slightly elevated skin temperature, prominent superficial veins, significant tenderness, and restricted knee joint movement. X-ray imaging demonstrates lytic bone destruction at the distal end of the femur, with the presence of Codman's triangle. | Given the patient's symptoms, physical examination findings, and X-ray results, the recommended imaging examination would be a Magnetic Resonance Imaging (MRI) of the right femur. The MRI will provide detailed images of the bone and surrounding soft tissues, which is essential for determining the extent of the lesion, any potential pathological fracture, soft tissue mass associated with the primary lesion, or possible invasion into joint space or adjacent neurovascular structures. | The main diagnosis to consider for this patient is osteosarcoma, a type of bone cancer that most often occurs in teenagers and young adults and is frequently located at the distal femur. Symptoms typically include pain at the tumor site that can become worse during the night or with activity, swelling and redness. The presence of lytic bone destruction and Codman's triangle on X-ray are classic signs of osteosarcoma. | The treatment plan for osteosarcoma generally consists of neoadjuvant chemotherapy to shrink the tumor, followed by surgical excision to remove the remaining tumor. Depending on the location and size of the tumor, limb salvage surgery may be possible, where the tumor is removed while preserving the affected limb. Postoperative chemotherapy is usually indicated to prevent metastasis and recurrence. Prosthetic or reconstructive surgery may also be needed depending on the extent of the tumor removal. |
| 46 | A 38-year-old female has been experiencing swollen and painful joints in both upper and lower extremities for the past 10 years. X-ray imaging reveals multiple areas of bone destruction in the finger joints and wrist joints. Despite joint examination, there are still multiple painful swollen joints. The spleen is palpable 2cm below the rib margin, with a moderate consistency. Laboratory results show a white blood cell count (WBC) of 2×109/L, a platelet count of 60×109/L, an erythrocyte sedimentation rate (ESR) of 70mm/h, and a negative urinalysis. The patient has been taking nonsteroidal anti-inflammatory drugs (NSAIDs) for the past 10 years. | Based on the patient's history of long-term joint swelling, pain, and the observation of multiple areas of bone destruction in her finger and wrist joints, further imaging studies are necessary to fully comprehend the extent of skeletal involvement. Therefore, I would recommend a Magnetic Resonance Imaging (MRI) scan. An MRI scans is capable of providing detailed images of both bones and soft tissues like cartilage, ligaments, and tendons. This will allow us to assess any potential damage to these structures, which might not be visible on X-ray. | From the laboratory results of low WBC count, low platelet count, elevated ESR, along with the clinical manifestations of chronic joint pain, bone destruction, and an enlarged spleen, this patient seems to be suffering from Rheumatoid Arthritis (RA), with the possibility of Felty's syndrome. Felty's syndrome is a rare complication of longstanding rheumatoid arthritis characterized by the triad of RA, splenomegaly, and neutropenia. | The treatment plan should aim at reducing pain, inflammation and the progression of joint damage. The NSAID usage can be continued for symptom relief but disease-modifying antirheumatic drugs (DMARDs) like Methotrexate should be considered early in the course of the disease to prevent severe joint damage. In case of Felty's syndrome, immunosuppressive therapy may also be needed. Regular follow-ups are essential to monitor disease progression and adjust treatment accordingly. Physical therapy could be beneficial too in maintaining joint flexibility. If conservative treatments fail and joint damage is severe, surgical intervention (like joint replacement) may be considered. |
| 47 | The patient, a 33-year-old male, was emergently admitted to the hospital due to an injury to his right forearm. He was diagnosed with a Colles' fracture of the right wrist. Manual reduction followed by plaster external fixation was applied. Six weeks later, after removal of the plaster, significant limitation in flexion and extension of the right wrist and all fingers was observed. | Given this patient's history and the limited movement of his right wrist and fingers after plaster removal, it's necessary to perform further imaging studies. An X-ray of the right forearm and wrist should be done initially to check for malunion or nonunion of the fractured bone, which could be causing the restricted mobility.  If the X-ray does not fully explain the limitation in flexion and extension, I would then recommend Magnetic Resonance Imaging (MRI) to inspect the soft tissues around the fracture site. This can provide a detailed view of ligaments, tendons, muscles, and nerves, which might have been affected during the initial injury or subsequent treatment. | The primary diagnosis appears to be post-traumatic stiffness of the wrist and fingers, which may have resulted from immobilization or possible damage to nerves, tendons, or ligaments. | The treatment plan should first focus on physical rehabilitation with a combination of active and passive exercises under the guidance of a physiotherapist. The goal is to gradually restore range of motion and function. Hand therapy, which focuses specifically on rehabilitation techniques for hand and arm conditions, could also be beneficial. |
| 48 | A 10-year-old male student presents with left knee pain that started 36 hours ago without any apparent cause. Initially, the pain was mild, and he continued to attend school for one day. However, in the afternoon after school, he limped back home. The pain worsened in the evening, causing him to avoid weight-bearing on the affected limb. He is unable to fully extend his leg while lying in bed, finding relief in a semi-flexed position. Additionally, he experiences generalized chilling and shivering, followed by the onset of fever. His body temperature has not been measured. | Given the symptoms of unilateral knee pain and swelling, inability to fully extend the leg, and recent onset fever, it is necessary to perform imaging studies on this young patient. My primary recommendation would be a plain X-ray of the knee to rule out any obvious bone abnormalities or trauma.  However, considering the rapid progression of symptoms, an MRI should also be considered for a more in-depth view of the joint's soft tissue structures. An MRI will help assess possible inflammation or infection in the surrounding tissues such as muscles, tendons, ligaments, and even the joint capsule itself. If infection is suspected, a joint fluid analysis may be required for definitive diagnosis. | Based on the sudden onset of symptoms without an apparent cause, the patient's age, and the other symptoms present (fever, chills), the primary diagnosis could be septic arthritis (infectious arthritis). This is a serious condition where a joint becomes infected with bacteria, resulting in painful inflammation. | This is a medical emergency and requires immediate attention. The treatment plan involves antibiotics to treat the underlying bacterial infection. These are usually given intravenously at first but can be switched to oral form once improvement is noted. Severe cases may require surgical drainage of the joint to remove the infected fluid.  In addition to treating the infection, management of septic arthritis also involves pain relief and maintaining joint function. This may involve physical therapy after the acute infection has been controlled. It is important to start this therapy early to prevent long-term stiffness and improve the likelihood of a complete recovery.  Regular follow-ups are essential to monitor progress and adjust treatment accordingly. If not treated promptly and appropriately, septic arthritis can lead to severe complications including irreversible damage to the joint. |
| 49 | A 22-year-old female presents with a progressive bending deformity at the upper end of the left thigh and shortening of the left lower limb over the past six months. X-ray imaging reveals curvature of the left femoral neck, thickening and expansion of the cortex, and thinning of the cortical bone. The medullary cavity demonstrates expansile lytic changes, resembling a ground-glass appearance. | Based on the description provided, the patient has already had an X-ray imaging test, which demonstrates some pathological changes. However, to gain a more detailed understanding of the condition including soft tissues involvement, a Magnetic Resonance Imaging (MRI) scan is recommended. This will provide a detailed image of the bones, joint and surrounding tissues such as muscles, ligaments, and tendons. Furthermore, it can detect any tumor or abnormal growth in the bone marrow and will be helpful in assessing the extent of the disease. | Considering the patient’s symptoms and X-ray findings, the primary diagnosis would likely be Fibrous Dysplasia. This is a bone disorder where normal bone is replaced with fibrous bone tissue resulting in deformities or fractures. It often occurs during childhood or adolescence and may affect one or many bones. The common site of involvement is the femur, tibia, skull, ribs, and pelvis. | In terms of treatment approach, management of fibrous dysplasia is primarily symptomatic. Medications could include pain relievers for discomfort and bisphosphonates to slow down bone turnover and decrease pain. Surgical intervention may be required depending on the severity of the disease. This could involve fracture repair, correction of deformities, or removal of affected bone and replacement with grafting.  Physical therapy may also be beneficial for muscle strengthening around the affected area and improving overall function. Regular follow-ups would be necessary for monitoring the progress of the disease. It is also important to note that treatment varies from patient to patient and depends on the location, size, and number of fibrous dysplastic areas. Therefore, an individualized treatment plan should be considered based on each patient's conditions. |
| 50 | A 60-year-old male patient has been experiencing left shoulder pain for over two months. The pain is clearly related to movement and posture, gradually worsening and extending to the middle part of the upper arm. Activities such as external rotation, abduction, and extension are significantly restricted due to the pain. Physical examination reveals mild atrophy of the left deltoid muscle, along with notable tender points in the biceps tendon, supraspinatus muscle, and infraspinatus muscle. There is also a significant limitation in shoulder joint mobility. | Based on the description provided, this patient should undergo X-ray of the left shoulder initially to evaluate for degenerative changes like arthritis or bony abnormalities. Following this, an MRI scan of the shoulder would be advantageous. The MRI can provide a detailed understanding of the soft tissue structures such as tendons, ligaments and muscles around the shoulder joint. This will help in identifying inflammation, tears or any other damage to these structures. | The symptoms presented by the patient including pain with movement and postural activities, decreased range of motion and tenderness around certain muscles suggest a diagnosis of Rotator Cuff Tendinitis or possible Rotator Cuff Tear. The rotator cuff is a group of muscles and tendons that surround the shoulder joint, providing stability and allowing a wide range of motion. Overuse or injury can cause inflammation or tears in these structures leading to pain and limited mobility. | In terms of treatment approach, initial management typically involves conservative measures such as rest, ice application, and non-steroidal anti-inflammatory drugs (NSAIDs) for pain relief. Physical therapy can also be beneficial, focusing on strengthening exercises for the shoulder muscles and maintaining joint mobility. Steroid injections may be considered if conservative treatment fails to alleviate the symptoms.  If the patient has a significant tear in the rotator cuff as determined by the MRI, surgical intervention may be necessary. This could involve arthroscopic surgery or open repair depending upon the severity of the tear. Post-surgical physical therapy would then be vital for restoring function and range of motion. Regular follow-up is crucial to monitor progress and adjust treatment plans accordingly. |
