## supplemental Table 2 for "Assessing the performance of GPT-4 in the filed of osteoarthritis and orthopaedic case consultation"

Supplementary Table 2: Questions and recommendations from China guideline of OA, response from GPT-4

|  | Question | Recommendation | GPT-4 response |
| --- | --- | --- | --- |
| 1 | What are the high-risk populations for osteoarthritis? | Recommendation 1: Individuals who have one or more of the following risk factors are considered high-risk populations for osteoarthritis (OA): age 40 years and above, female gender, obesity or overweight, history of trauma (strong recommendation, evidence level: B).  Recommendation 2: High-risk populations for knee osteoarthritis (OA) also include individuals with periarticular muscle atrophy, long-term engagement in heavy labor or specific occupations, family history of OA, residing in high-risk areas, or having gut dysbiosis (weak recommendation, evidence level: C).  Recommendation 3: Individuals at high risk of hip osteoarthritis (OA) also include those with the presence of risk factors such as hip dysplasia, cam deformity of the femoral neck, long-term engagement in weight-bearing labor or other specific occupations, and a family history of OA patients (Recommendation strength: Strongly recommend, Evidence level: C).  Recommendation 4: Individuals at high risk of hand OA also include those with the presence of risk factors such as long-term engagement in specific hand labor, being in perimenopause, a family history of OA patients, or dysbiosis of gut microbiota (Recommendation strength: Weakly recommend, Evidence level: C). | Osteoarthritis (OA) is a degenerative joint disease, and certain populations are at a higher risk than others. These include:  1. **Older adults**: The risk of developing OA increases with age. Most people over the age of 60 have OA to some degree, but its severity varies.  2. **Women**: Women are more likely than men to develop OA, especially after the age of 50.  3. **Obese individuals**: Extra body weight puts additional pressure on weight-bearing joints like the knees and hips, increasing the risk of OA. Also, fat tissue produces proteins that may cause harmful inflammation in and around the joints.  4. **Individuals with joint injuries**: People who have had a joint injury, such as a fracture or ligament tear, are more likely to eventually develop OA in that joint.  5. **People with genetic predisposition**: Some people inherit genes that make them more likely to develop OA. For instance, individuals with familial forms of hand OA or hip OA have genetic mutations that cause the condition.  6. **People with certain occupations**: Jobs that involve repetitive stress on a particular joint can increase the risk of developing OA in that joint. For example, professional athletes, construction workers, or those who do heavy lifting as part of their jobs.  7. **People with certain metabolic or endocrine disorders**: Conditions like diabetes and hypothyroidism can also increase the risk of developing OA.  8. **Individuals with other joint diseases or malformations**: People who have rheumatoid arthritis, gout, or congenital joint malformations are also at a higher risk.  Remember that while these factors can increase the risk, they don't guarantee that someone will develop OA. Many people with one or more risk factors never develop the disease, while others with no known risk factors do. It's also important to note that although OA is often associated with aging, it's not a natural part of aging, and healthy lifestyle choices can reduce the risk and slow progression if it does occur. |
| 2 | What are the main clinical manifestations of osteoarthritis? | Recommendation 5:Joint pain and limited joint mobility are the most common clinical symptoms of osteoarthritis (OA) (strong recommendation, evidence level: C).  Recommendation 6: Tenderness and joint deformity are the most common signs of hand osteoarthritis (OA) and knee OA. Crepitus (sensation) and muscle atrophy are commonly observed in knee OA (Recommendation strength: Strongly recommend, Evidence level: C). | Osteoarthritis (OA) is a degenerative joint disease that affects the whole joint including bone, cartilage, ligaments, and muscles. Its clinical manifestations can vary, but the most common signs and symptoms include:  1. **Pain**: This is typically the first symptom of OA. The affected joint may hurt during or after movement.  2. **Stiffness**: Joint stiffness is most noticeable upon waking up in the morning or after a period of inactivity.  3. **Loss of flexibility**: Individuals may not be able to move their joint through its full range of motion.  4. **Crepitus**: This is a grating or popping sensation or noise when the joint is used.  5. **Swelling**: This can occur because of inflammation within the joint or the production of extra synovial fluid, a natural lubricant in your joints.  6. **Bone spurs**: These are extra bits of bone, which feel like hard lumps, that form around the affected joint.  7. **Muscle weakness**: Particularly in OA of the knee, the muscles surrounding the joint may become weaker.  8. **Functional impairment**: As the disease progresses, it might become difficult to perform daily activities that require the use of the joint, such as walking, climbing stairs, opening jars, or typing.  The intensity of these symptoms can vary from person to person, and even from day to day for the same person. Also, it's important to remember that the severity of symptoms might not directly correspond with the degree of joint damage — some people experience severe symptoms with minor damage, while others have little to no symptoms despite severe damage.  If you suspect you have OA or are experiencing worsening symptoms of existing OA, please seek medical attention. It's a chronic condition, but several treatment options can help manage the symptoms and improve quality of life. |
| 3 | What imaging examinations should clinical physicians select for suspected osteoarthritis patients? | Recommendation 7:X-ray examination is the preferred choice for suspected osteoarthritis (OA) patients. If necessary, further examinations such as CT, MRI, and ultrasound can be performed to determine the location and extent of degeneration and facilitate differential diagnosis (strong recommendation, evidence level: B). | Imaging is an essential part of the diagnostic process for osteoarthritis (OA). The type of imaging depends on the suspected location of the OA, the severity of the patient's symptoms, and what information the doctor is trying to gather. Here are some common types of imaging used:  1. **X-ray**: This is the most commonly used imaging technique for diagnosing OA. It can show bone damage, cartilage loss (seen as a narrowing of the space between the bones), bone spurs, and changes in the shape or alignment of the joints.  2. **MRI (Magnetic Resonance Imaging)**: While not typically the first choice, an MRI can provide a more detailed image and show damage to other structures within the joint like ligaments and tendons, in addition to the bones and cartilage. MRI might be more helpful in the early stages of the disease when X-ray findings are not yet significant, or when more information is needed about the condition of the joint.  3. **Ultrasound**: This is less commonly used for OA, but it can be helpful in evaluating soft tissue structures around the joint, such as bursae, and can also reveal fluid within the joint or bone spurs.  4. **CT scan (Computed Tomography)**: While not commonly used for routine evaluation of OA, it can be helpful in some cases to assess the bony anatomy of the joint, particularly before joint replacement surgery.  In addition to imaging studies, the clinical diagnosis of OA often involves a medical history assessment, physical examination, and sometimes laboratory tests to help rule out other diseases. The choice of imaging will be guided by the physician based on the individual patient's circumstances. |
| 4 | What diseases should be distinguished from osteoarthritis during clinical diagnosis? | Recommendation 8: Clinical physicians should differentiate osteoarthritis (OA) from other diseases that can cause joint pain and functional impairment, including autoimmune arthritis, infectious arthritis, gout, pseudogout, and joint injuries (GPS). | During the diagnostic process for osteoarthritis (OA), it's important to differentiate it from other conditions that can cause similar symptoms, such as joint pain and stiffness. Here are some conditions that should be considered:  1. **Rheumatoid Arthritis (RA)**: This is an autoimmune disorder that typically affects joints symmetrically (for example, both knees or both hands), while OA often affects joints asymmetrically. RA also has systemic symptoms like general fatigue and feeling unwell, which are not typical of OA.  2. **Gout**: This is caused by the deposition of uric acid crystals in the joint, typically the big toe. Attacks of gout can cause severe pain and swelling in the affected joint, and while gout can cause OA over time, it requires a different initial treatment approach.  3. **Lupus**: This is an autoimmune disease that can cause a variety of symptoms, including joint pain, skin rashes, kidney problems, and others. It requires a different treatment approach than OA.  4. **Psoriatic Arthritis**: This typically occurs in people with psoriasis and can affect any joint in the body. Psoriatic arthritis can cause symptoms of inflammation like warmth and swelling in the joints, which is less common in OA.  5. **Fibromyalgia**: This is a chronic pain syndrome that can cause widespread pain and tenderness. People with fibromyalgia often have normal joint examinations, unlike those with OA, where physical findings can show joint changes.  6. **Infectious Arthritis (Septic Arthritis)**: This is caused by an infection in the joint. It typically causes severe pain, swelling, and redness, and can cause rapid damage to the joint if not treated promptly.  7. **Reactive Arthritis**: This is a form of arthritis that occurs as a reaction to an infection elsewhere in the body.  8. **Other degenerative joint conditions**: Conditions like avascular necrosis or hip dysplasia can cause joint pain and have radiographic changes that might mimic OA.  9. **Osteoporosis**: While osteoporosis itself does not cause joint pain, fractures due to osteoporosis can cause pain and might be mistaken for OA.  These conditions often require different treatments than OA, so correct diagnosis is crucial for effective management. As always, if a person suspects they have OA or another joint condition, they should consult a healthcare provider for evaluation. |
| 5 | What are the treatment goals and principles for osteoarthritis？ | Recommendation 9: Clinical physicians should adopt a stepped and individualized treatment approach based on the patient's age, gender, body mass index (BMI), location and severity of lesions, aiming to reduce pain, improve or restore joint function, enhance the patient's quality of life, delay disease progression, and correct deformities (GPS). | Osteoarthritis (OA) is a chronic degenerative disease that cannot be cured, so treatment goals and principles focus on managing symptoms and improving quality of life. Here are the main treatment goals for OA:  1. **Pain management**: The most immediate goal is usually to reduce pain to a manageable level.  2. **Maintain/improve joint function**: This involves maintaining or improving the range of motion of the joint and strength of the surrounding muscles.  3. **Minimize disability**: This goal is to keep the person as mobile and functional as possible to enable them to carry out their daily activities.  4. **Slow disease progression**: While we can't stop OA, certain strategies can help slow down its progression.  5. **Educate and involve the patient in the management of their disease**: OA requires long-term self-management, so it's important to educate patients about the disease and involve them in the management process.  It's important to remember that treatment plans are individualized, based on the person's age, overall health, severity and location of OA, and personal preferences. A combination of treatments is often used, and it might take some trial and error to find what works best for each individual. |
| 6 | What is the basic treatment of osteoarthritis ? | Recommendation 10: It is strongly recommended to prioritize basic treatment options for patients with osteoarthritis (OA), including health education, exercise therapy, physical therapy, and assistive support (strong recommendation, evidence level: B). | The basic treatment for osteoarthritis (OA) typically involves a combination of non-pharmacological and pharmacological strategies designed to relieve symptoms, improve joint function, and slow disease progression. Here are the fundamental aspects of OA treatment:  1. **Non-pharmacological treatments:**  - **Exercise**: Regular physical activity helps maintain joint mobility and strength, reduces pain, and improves overall health. Both aerobic activities (like walking, biking, or swimming) and strength training can be beneficial. A physical therapist can provide a personalized exercise plan.    - **Weight management**: If a patient is overweight, losing weight can reduce stress on weight-bearing joints and decrease pain. A healthy diet and regular exercise are key components of weight management.    - **Physical therapy**: Physical therapists can provide treatments to relieve pain, improve movement, and strengthen muscles around affected joints. They might also recommend assistive devices, such as braces, shoe inserts, or canes, to help take pressure off affected joints and improve mobility.    - **Patient education and self-management programs**: Understanding the disease and learning how to manage it can help improve quality of life and slow the progression of OA.  2. **Pharmacological treatments:**  - **Pain relievers**: Over-the-counter (OTC) pain relievers like acetaminophen (Tylenol) or nonsteroidal anti-inflammatory drugs (NSAIDs) like ibuprofen (Advil, Motrin) are often the first line of treatment for OA pain. Topical creams or gels containing NSAIDs or capsaicin might also be used.  - **Prescription medications**: If OTC medications aren't sufficient, stronger medications may be prescribed, such as prescription NSAIDs, tramadol, or in some cases, opioids. However, opioids are generally considered a last resort due to their risk of dependency and other side effects.  3. **Complementary and alternative therapies:** Some people with OA find relief with alternative therapies, such as acupuncture, massage, tai chi, or yoga. However, the effectiveness of these treatments can vary from person to person.  These are the basic treatment strategies for OA. For more advanced disease or in cases where these treatments aren't effective, other options like injections or surgery might be considered. Treatment is individualized to the person's symptoms, severity of disease, overall health, and personal preferences. As always, treatment decisions should be made in collaboration with a healthcare provider. |
| 7 | What is the efficacy of exercise therapy for osteoarthritis? | Recommendation 11: Aerobic exercise and aquatic exercise have been shown to improve pain and function in patients with knee and hip osteoarthritis. It is recommended that clinicians develop individualized exercise therapy plans based on the patient's condition (Recommendation strength: strong recommendation, Evidence grade: B).  Recommendation 12: Hand exercises can alleviate pain and joint stiffness in patients with hand osteoarthritis (OA). It is recommended that patients with hand OA engage in hand exercises (Recommendation strength: Strongly recommend, Evidence level: C). | Exercise therapy is considered a cornerstone of osteoarthritis (OA) management. Numerous studies have shown its efficacy in reducing pain, improving physical function, and enhancing quality of life for individuals with OA. Here's how exercise therapy can benefit those with OA:  1. **Pain reduction**: Regular exercise has been shown to decrease pain in individuals with OA. The exact mechanisms for this pain reduction are not entirely understood, but it's thought that exercise may help by increasing blood flow to the joints, strengthening the muscles that support the joints, and releasing natural pain-relieving endorphins.  2. **Improved physical function**: Exercise can help improve flexibility, strength, and balance, all of which can help increase range of motion and joint stability, making daily activities easier to perform. This can be especially important for individuals with OA, who often experience stiffness and decreased joint function.  3. **Weight management**: If a person is overweight or obese, exercise can help manage weight, which reduces pressure on weight-bearing joints and can decrease pain.  4. **Better mood and quality of life**: Regular physical activity is known to have benefits for mental health, including reduced symptoms of depression and anxiety. This can be particularly beneficial for people with chronic conditions like OA, as pain and physical limitations can often lead to psychological distress.  5. **Potentially slow disease progression**: While more research is needed in this area, some studies suggest that regular exercise may help slow the progression of OA by maintaining healthy joint cartilage and delaying further joint damage.  Different types of exercises can be beneficial, including:  - **Aerobic exercise**: Low-impact activities like walking, cycling, swimming, and water aerobics can improve cardiovascular health without putting undue stress on the joints.  - **Strength training**: Strengthening the muscles that support the affected joint can help improve stability and reduce pain.  - **Flexibility and balance exercises**: These can help maintain joint mobility, reduce stiffness, and prevent falls.  Before starting an exercise program, it's important for those with OA to talk with their healthcare provider or a physical therapist. They can provide guidance on which types of exercises are safe and appropriate based on the person's condition, overall health, and physical capabilities. They can also help ensure the exercises are being done correctly to avoid injury and get the most benefit. As with any treatment, individual responses can vary. |
| 8 | What is the effectiveness of physical therapy for osteoarthritis? | Recommendation 13: Interferential current electrical stimulation and pulsed ultrasound therapy may be considered for relieving pain symptoms in patients with knee osteoarthritis (Recommendation strength: weak recommendation, Evidence grade: B).  Recommendation 14: Hydrotherapy, cryotherapy, thermotherapy, mud bath therapy, radiofrequency ablation, and other transcutaneous electrical nerve stimulation methods have shown some effectiveness in the treatment of osteoarthritis. However, there is currently a lack of standardized protocols. Clinicians may use these therapies at their discretion (Recommendation strength: weak recommendation, Evidence grade: C). | Physical therapy is widely recognized as an effective treatment for managing the symptoms of osteoarthritis (OA). The goal of physical therapy in OA is to reduce pain, improve mobility, increase strength, and enhance the ability to do everyday activities. Here's how physical therapy can help those with OA:  1. **Pain reduction**: Physical therapists can use a variety of techniques to help reduce pain, including therapeutic exercises, manual therapy (like massage and joint manipulation), and modalities like heat, cold, ultrasound, or electrical stimulation.  2. **Improved physical function**: Therapists provide exercises designed to improve range of motion, flexibility, balance, coordination, and strength, all of which can help individuals with OA move better and do their daily activities more easily.  3. **Strength training**: Strengthening exercises target muscles that support the affected joints, which can help relieve pain and improve function.  4. **Education**: Physical therapists can provide education on ways to manage symptoms, joint protection techniques, the use of assistive devices (like canes or shoe inserts), and the importance of maintaining a healthy lifestyle, including regular physical activity and a balanced diet.  5. **Weight management**: For individuals who are overweight, physical therapists can provide guidance on safe exercises to help with weight loss, which can help reduce stress on weight-bearing joints.  6. **Gait analysis and correction**: If OA is affecting the lower extremities, therapists can analyze a person's gait and suggest corrections to reduce strain on the joints when walking.  Research supports the effectiveness of physical therapy for OA. A systematic review published in the Journal of Rheumatology in 2015 concluded that exercise programs delivered by physical therapists provide both short-term and long-term benefits for people with knee OA, including reduced knee pain and improved physical function and quality of life.  As with any treatment, individual responses to physical therapy can vary. It's important for the physical therapist and patient to communicate regularly about the effectiveness of the treatment and make adjustments as needed. It's also important to remember that successful physical therapy requires active participation from the patient, including doing prescribed exercises at home and making lifestyle changes as recommended. |
| 9 | How should pharmacological treatment for osteoarthritis be selected? | Recommendation 15: It is strongly recommended to use topical non-steroidal anti-inflammatory drugs (NSAIDs) as first-line treatment for pain in patients with knee osteoarthritis, especially those with coexisting gastrointestinal, cardiovascular diseases, or frailty (Recommendation strength: strong recommendation, Evidence grade: A).  Recommendation 16: It is recommended for patients with persistent or moderate to severe pain in osteoarthritis to choose oral NSAIDs, including non-selective NSAIDs and selective cyclooxygenase-2 (COX-2) inhibitors, but caution should be taken regarding gastrointestinal and cardiovascular adverse events (Recommendation strength: strong recommendation, Evidence grade: B).  Recommendation 17: It is not recommended to use strong opioids for pain management in osteoarthritis, and cautious use of weak opioids such as tramadol is advised (Recommendation strength: strong recommendation, Evidence grade: C).  Recommendation 18: For patients with long-term, chronic, widespread pain and/or comorbid depression in osteoarthritis, the use of anxiolytic medications such as duloxetine is recommended (Recommendation strength: strong recommendation, Evidence grade: B). | Pharmacological treatment for osteoarthritis (OA) is typically selected based on several factors, including the severity of the patient's symptoms, their overall health status, the presence of any other medical conditions, potential side effects of the medication, and the patient's personal preferences.  1. **Over-the-Counter (OTC) Medications**: For mild to moderate OA pain, over-the-counter pain relievers like acetaminophen (Tylenol) or nonsteroidal anti-inflammatory drugs (NSAIDs) like ibuprofen (Advil, Motrin) are often the first line of treatment. Topical creams or gels containing NSAIDs or capsaicin may also be used.  2. **Prescription Medications**: If OTC medications aren't sufficient, stronger medications may be prescribed, such as prescription-strength NSAIDs, COX-2 inhibitors like celecoxib (Celebrex), tramadol, or in some cases, opioids. However, opioids are generally considered a last resort due to their risk of dependency and other side effects.  3. **Injections**: If oral medications are not enough, injections of corticosteroids or hyaluronic acid into the affected joint might be considered. These can provide temporary relief from symptoms.  4. **Topical Agents**: For some patients, particularly those who cannot tolerate oral NSAIDs due to gastrointestinal issues, topical NSAIDs can be a good option.  The potential benefits of each medication need to be weighed against the potential risks. For example, while NSAIDs can be effective in managing OA pain, they also come with risks, such as gastrointestinal bleeding, cardiovascular problems, and kidney damage, especially with long-term use or in individuals with existing health problems. |
| 10 | What is the efficacy and safety of intra-articular drug therapy for treating osteoarthritis? | Recommendation 19: Clinicians should exercise caution when using intra-articular corticosteroid injections for the treatment of osteoarthritis. Although they can provide rapid pain relief and improve joint function, there is a risk of accelerating cartilage loss with long-term and repeated use (Recommendation strength: strong recommendation, Evidence grade: B).  Recommendation 20: Clinicians may consider using intra-articular hyaluronic acid injections for the treatment of osteoarthritis. It can provide short-term pain relief, improve joint function, and reduce the need for analgesic medications, with a higher level of safety (Recommendation strength: weak recommendation, Evidence grade: B). | Intra-articular drug therapy, which involves injecting medications directly into the joint, is a common treatment option for osteoarthritis (OA), particularly for patients who haven't responded well to oral medications or have significant side effects from these medications. The most commonly used intra-articular therapies for OA are corticosteroids and hyaluronic acid.  **Corticosteroids**: Corticosteroids (such as triamcinolone or methylprednisolone) have strong anti-inflammatory effects and can provide rapid, short-term relief of OA pain. A review of studies published in the Cochrane Database of Systematic Reviews in 2015 found that intra-articular corticosteroids improved pain in people with knee OA for up to two weeks, but the effect diminished after that.  The main risks of corticosteroid injections are joint infection and damage to the joint tissue, particularly with frequent injections. There can also be temporary pain and swelling in the joint after the injection. Systemic side effects (like elevated blood sugar) can occur but are less common than with oral corticosteroids.  **Hyaluronic Acid**: Hyaluronic acid is a substance that is naturally present in the joint fluid and helps lubricate and cushion the joint. In people with OA, the quality and quantity of hyaluronic acid can be diminished, so injecting it into the joint can help improve joint function and reduce pain.  A 2012 meta-analysis published in the Annals of Internal Medicine found that hyaluronic acid injections provided a small to moderate improvement in pain and function in people with knee OA, but the clinical significance of these effects was uncertain.  As with corticosteroid injections, the main risks of hyaluronic acid injections are joint infection and joint damage. Pain and swelling at the injection site can also occur.  In general, while intra-articular therapies can provide relief from OA symptoms, they do not slow the progression of the disease. Therefore, they are typically used in conjunction with other treatments, such as oral medications, exercise, and physical therapy.  As with any treatment, the decision to use intra-articular therapy should be made through shared decision-making between the patient and their doctor, taking into account the severity of the patient's symptoms, the potential benefits and risks of the treatment, the patient's other health conditions and medications, and the patient's personal preferences. |
| 11 | What is the efficacy and safety of traditional Chinese medicine (TCM) treatment for osteoarthritis? | Recommendation 21: Topical and oral traditional Chinese medicine (TCM) can provide pain relief and improve joint function in osteoarthritis, with a higher level of safety. Clinicians may consider using them, but caution should still be taken to prevent skin allergies when using topical applications (Recommendation strength: weak recommendation, Evidence grade: B).  Recommendation 22: Acupuncture has been found to effectively improve joint pain and function in patients with osteoarthritis, and it is considered safe. It may be used at the clinician's discretion for the treatment of osteoarthritis (Recommendation strength: weak recommendation, Evidence grade: B). | Traditional Chinese Medicine (TCM) has been used for centuries to treat a variety of conditions, including osteoarthritis (OA). TCM approaches to OA often include herbal remedies, acupuncture, Tai Chi, and other mind-body practices. However, while many patients report relief from these treatments, the scientific evidence varies, and the quality of research studies can be inconsistent.  1. **Herbal remedies**: Many different herbs are used in TCM for OA, often in complex mixtures. Some studies suggest certain herbs may have anti-inflammatory and pain-relieving effects, but more high-quality research is needed to confirm these findings and to assess their safety, particularly given the risks of interactions with other medications and potential side effects.  2. **Acupuncture**: This technique, which involves inserting thin needles at specific points on the body, is often used in TCM to treat pain. Some studies suggest acupuncture may provide some pain relief for OA, but results have been mixed. A systematic review published in the British Journal of Sports Medicine in 2020 found that acupuncture seemed to slightly reduce pain and improve function in people with OA, but the researchers noted that the effects were small and may not be clinically relevant. The safety of acupuncture is generally considered to be good when performed by a competent practitioner, but potential side effects include infection, bleeding, or injury from needles.  3. **Tai Chi**: This mind-body practice, which involves slow, gentle movements and deep breathing, can help improve balance, strength, and flexibility. Several studies have found that Tai Chi can help reduce pain and improve physical function in people with knee OA. A systematic review and meta-analysis published in the Journal of Physiotherapy in 2019 concluded that Tai Chi may improve pain and physical function in people with OA. The safety of Tai Chi is generally considered good, but people with certain health conditions (like severe osteoporosis or heart problems) should check with their healthcare provider before starting Tai Chi.  It's important to note that TCM should not replace conventional medical treatments for OA, but can be used as a complementary approach. Before starting any new treatment, including TCM, it's important to talk with a healthcare provider. The provider can provide information about potential risks and benefits and ensure the treatment is appropriate given the patient's overall health and treatment plan. |
| 12 | What are the indications for arthroscopic surgery in the treatment of osteoarthritis? | Recommendation 23: Arthroscopic surgery for the treatment of knee osteoarthritis is only short-term effective in relieving pain symptoms, and there is no significant difference in medium to long-term efficacy compared to conservative treatments. Clinicians may consider it on a case-by-case basis (Recommendation strength: strong recommendation, Evidence grade: A).  Recommendation 24: Arthroscopic debridement is recommended for the treatment of knee osteoarthritis with locking symptoms (Recommendation strength: strong recommendation, Evidence grade: B).  Recommendation 25: For patients with shoulder osteoarthritis who are not suitable for shoulder joint replacement surgery due to age, activity level, or personal preference after other interventions have failed, arthroscopic debridement may be considered to improve symptoms (Recommendation strength: weak recommendation, Evidence grade: D). | Arthroscopic surgery involves inserting a small camera, called an arthroscope, into the joint through a small incision to visualize, diagnose, and treat joint problems. While it has been commonly used for many joint conditions, its use in treating osteoarthritis (OA) is a matter of ongoing debate.  Arthroscopy can be used for OA to remove loose fragments of cartilage, repair torn meniscus, or smooth out rough surfaces of joints. However, it's generally not considered a first-line treatment and is usually reserved for specific cases.  Guidelines published by various medical societies in recent years, such as the American Academy of Orthopaedic Surgeons (AAOS) and the Osteoarthritis Research Society International (OARSI), generally discourage the use of arthroscopy for degenerative knee disease (which includes OA), unless there are specific mechanical symptoms such as locking, catching, or instability that might be improved with surgery.  Several studies and reviews, including a significant study published in the New England Journal of Medicine, have shown that arthroscopic surgery does not lead to any substantial benefits for most people with knee OA. In fact, in some cases, it may cause more harm than good due to the risks associated with surgery, such as infection or complications from anesthesia.  However, every patient is unique, and the decision to proceed with arthroscopy should be made on a case-by-case basis, taking into account the specific characteristics of the patient's disease, their overall health, and their personal preferences. The decision should be made in consultation with a knowledgeable orthopedic surgeon, and the patient should be fully informed about the potential benefits and risks.  Lastly, remember that while surgery can help manage the symptoms of OA, it doesn't cure the disease. Even after surgery, patients will still need to engage in other treatment strategies such as physical therapy, weight management, and possibly medications to manage symptoms and prevent further joint damage. |
| 13 | What are the indications for osteotomy in the treatment of osteoarthritis? | Recommendation 26: For patients with single-compartment osteoarthritis of the tibiofemoral joint, especially in younger individuals with high activity levels and poor alignment, clinicians may consider osteotomy procedures such as high tibial osteotomy, distal femoral osteotomy, or proximal fibular osteotomy to improve joint function and alleviate pain (Recommendation strength: weak recommendation, Evidence grade: C).  Recommendation 27: For patients with mild hip osteoarthritis due to developmental dysplasia of the acetabulum, clinicians may consider acetabular osteotomy on a case-by-case basis (Recommendation strength: weak recommendation, Evidence grade: D). | Osteotomy is a surgical procedure that involves cutting and reshaping of bone. It is considered for the treatment of osteoarthritis in certain cases, with key indications including:  1. **Younger Age**: Osteotomy is often considered for younger patients who have arthritis, typically those under 60-65 years of age. This is because joint replacement surgeries, although effective, may wear out over time and may not be suitable for younger, more active individuals.  2. **Unicompartmental (one-sided) Osteoarthritis**: If the arthritis only affects one side of the knee (medial or lateral compartment), an osteotomy may help shift the body's weight off the damaged side.  3. **Early/Moderate Disease**: Osteotomy is usually considered in the early to moderate stages of osteoarthritis, before the joint damage becomes too extensive.  4. **Realignment Need**: Osteotomy can be useful for realigning a joint that has become deformed due to arthritis.  5. **Active Lifestyle**: For people who participate in high-impact activities, osteotomy might be a better option as compared to joint replacement which may be subject to faster wear and tear.  It's important to note that while osteotomy can effectively relieve pain and improve function in selected patients, it doesn't stop the progression of arthritis. Therefore, patients may still require a joint replacement in the future. As always, the decision to proceed with osteotomy should be made in consultation with a healthcare provider. |
| 14 | What are the indications for total joint replacement surgery? | Recommendation 28: Total joint replacement surgery is recommended for patients with severe osteoarthritis who have not responded to other interventions. Factors such as the patient's specific condition, subjective preferences, and expectations should also be taken into consideration (Recommendation strength: strong recommendation, Evidence grade: B). | Total joint replacement surgery, also known as arthroplasty, is a surgical procedure where the worn-out or damaged surfaces of the joint are removed and replaced with artificial parts. The most common joints replaced are the hip and knee. Here are the typical indications for total joint replacement surgery:  1. **Severe Pain**: When pain in the affected joint is severe enough to affect daily life activities such as walking, climbing stairs, or even resting, and when this pain isn't relieved by non-surgical treatments.  2. **Joint Stiffness**: Significant reduction in the range of motion of the joint that impedes routine tasks or activities.  3. **Moderate to Severe Arthritis**: This could be osteoarthritis, rheumatoid arthritis, or post-traumatic arthritis that has not responded well to conservative management.  4. **Joint Deformity**: Visible deformation of the joint may warrant joint replacement.  5. **Failed previous joint surgery**: If previous joint-preserving surgery has failed to relieve symptoms or if a previous joint replacement has failed.  6. **Poor Quality of Life**: Overall quality of life is significantly impacted due to constant pain and reduced joint mobility.  7. **Bone Damage**: X-rays or other imaging tests shows advanced arthritis or significant bone damage.  The decision for total joint replacement should be a mutual one between the patient and doctor, taking into account the patient's age, overall health, bone density, and the impact of symptoms on their quality of life. A thorough discussion of the risks and benefits of surgery, as well as potential alternatives, should always precede the decision to proceed with joint replacement surgery. |
| 15 | What are the indications for partial joint replacement surgery? | Recommendation 29: For patients with isolated patellofemoral osteoarthritis who have not responded well to other treatments, patellofemoral joint replacement surgery is recommended (Recommendation strength: strong recommendation, Evidence grade: C).  Recommendation 30: For patients with symptomatic single-compartment tibiofemoral osteoarthritis who meet the indications, unicompartmental knee replacement surgery is recommended (Recommendation strength: strong recommendation, Evidence grade: C). | Partial joint replacement surgery, also known as unicompartmental or hemiarthroplasty, is a surgical procedure where only the damaged part of the joint is replaced with an artificial implant, preserving the remaining healthy parts. This procedure is commonly performed on knees and hips. Here are some key indications for partial joint replacement surgery:  1. **Unicompartmental Disease**: The primary indication for partial joint replacement is when arthritis or damage is confined to one compartment (or section) of the joint. For instance, in the knee, if only the medial (inner) or lateral (outer) compartment is affected, a partial knee replacement may be considered.  2. **Preservation of Healthy Tissue**: When a significant amount of the joint tissue is still healthy, a partial replacement may be preferred to preserve as much of the natural joint as possible.  3. **Age** and **Activity Level**: Partial joint replacement can sometimes be suitable for younger, more active patients because it may preserve more joint function and feel more natural than a total replacement. It also allows for easier conversion to a total joint replacement later, if necessary.  4. **Less Severe Symptoms**: If symptoms do not significantly impair the patient's quality of life and are less severe, a partial replacement may be adequate to alleviate discomfort.  5. **Bone Quality**: Adequate bone quality is needed to support the implant. This makes partial replacements more suitable for those without extensive bone loss or deformities.  It's important to remember that each patient's situation is unique, and a thorough discussion with a healthcare provider is essential to determine the best treatment option. Factors such as overall health, lifestyle, disease progression, and personal preferences should all be taken into account when considering joint replacement surgery. |
